# *PD-L1*–linked spatial decoupling of tumour–immune interactions in EBV-positive DLBCL

**DOI:** 10.64898/2026.07.02.26357123

**Authors:** Axel Künstner, Matthias Kümmel, Anke Fähnrich, Stefanie Derer, Annika Raschdorf, Hanno M. Witte, Yamil Maluje, Benedikt Färber, Thies Rösner, Lisa Michelson, Friederike Stiller, Mona Maria Merz, Veronica Bernard, Stephanie Stölting, Daniel Kolbe, Wolfgang Peter, Hartmut Merz, Nikolas von Bubnoff, Alfred C. Feller, Hauke Busch, Philipp Lohneis, Niklas Gebauer

**Affiliations:** University of Luebeck, Medical Systems Biology Group, University of Lübeck, Ratzeburger Allee 160, 23538 Lübeck, Germany; University of Luebeck, Department of Hematology and Oncology, University Cancer Center Schleswig-Holstein (UCCSH) and University Hospital Schleswig-Holstein (UKSH), Campus Luebeck, Luebeck, Germany; University of Luebeck, Institute of Nutritional Medicine, University Hospital Schleswig-Holstein, Campus Lübeck, Lübeck, Germany; Department of Internal Medicine III Hematology and Oncology, University Hospital Ulm, Ulm, Germany; Hämatopathologie Lübeck, Reference Centre for Lymph Node Pathology and Hematopathology, Lübeck, Germany; Institute of Clinical Molecular Biology, Kiel University, University Hospital Schleswig-Holstein, Kiel, Germany; HLA Typing Laboratory of the Stefan-Morsch-Foundation, 557565 Birkenfeld, Germany

**Author notes:** **Corresponding Author:** Prof. Dr. med. Niklas Gebauer, Department of Haematology and Oncology, UKSH Campus Luebeck, Ratzenburger Allee 160, 23538 Luebeck. Shared first authorship (Axel Künstner, Matthias Kümmel). Shared senior authorship (Hauke Busch, Philipp Lohneis, Niklas Gebauer).

**Keywords:** DLBCL, EBV, Genomic landscape, PD-L1, spatial architecture

## Abstract

EBV-positive diffuse large B-cell lymphoma (DLBCL) is an aggressive lymphoma enriched in elderly patients, in which *PD-L1* genomic gains cooperate with viral signalling to promote immune escape. However, the spatial mechanisms by which *PD-L1* reshapes the tumour microenvironment remain unresolved.

Here, we integrate genomic profiling with spatial transcriptomics, spatial proteomics, and single-cell RNA sequencing to resolve the tumour–immune architecture of EBV-positive DLBCL. *PD-L1* gain tumours exhibit a striking decoupling of immune proximity and immune engagement: T cells accumulate in close proximity to tumour cells yet direct tumour–immune contact is largely absent. Tumour boundaries are enriched for cancer-associated fibroblasts and display transcriptional signatures of metabolic immune suppression, while tumour-proximal T cells exhibit pronounced exhaustion.

Together, these findings reveal a *PD-L1*–driven spatial immune evasion architecture that permits immune infiltration while enforcing multilayered suppression through fibroblast-mediated exclusion, metabolic constraint, and T-cell dysfunction, ultimately preventing effective anti-tumour immunity in EBV-positive DLBCL.

## Introduction

Epstein–Barr virus transforms B cells through a suite of latent proteins that simultaneously drive proliferation and enforce immune escape. Diffuse large B-cell lymphoma (DLBCL), the most common type of non-Hodgkin lymphoma (NHL), accounts for 30 – 40% of newly diagnosed lymphomas, of which ∼ 8% are EBV-positive corresponding to ∼ 27,000 cases and ∼13,300 deaths annually worldwide, and is more common in immunocompromised individuals and geographically in Africa, Asia, and Latin America ^1–7^. The disease predominantly affects older individuals (∼70 years) and often presents extranodally with poorer prognosis and a male predominance (1.4:1) ^6,8–11^. Although historically linked to poor outcomes, the prognostic impact of EBV has been attenuated in the rituximab era ^9,12–16^.

EBV-positive DLBCL displays polymorphic or monomorphic histology, expresses post–germinal centre B-cell markers, and is defined by EBER and LMP1 positivity with predominantly latency II/III programs ^7–9,12,15–22^. It represents a distinct molecular entity shaped by viral–host interactions and is typically characterized by an activated B-cell phenotype with enhanced NFκB signaling^15,23,24^. Recurrent alterations affect JAK–STAT signalling, chromatin remodelling, and immune evasion pathways, while only a minority of cases align with established molecular clusters, underscoring a distinct mutational architecture ^15,24–29^.

EBV latent proteins, including LMP1 and LMP2A, promote proliferation, NFκB activation, and immune escape. These effects are reinforced by PD-L1 overexpression, often driven by 9p24.1 amplification (∼ 30% of cases), alongside upregulation of immunosuppressive cytokines and impaired antigen presentation ^28,30–39^. The tumour microenvironment is further characterized by T-cell exhaustion (e.g., LAG3, TIM3) and a shift toward M2 macrophage polarization^31,40–45^. However, the mechanistic interplay between EBV latency programs and the immunosuppressive tumour microenvironment remains undefined at spatial and functional resolution, particularly given the substantial heterogeneity of EBV-positive DLBCL.

Here, we integrate whole-exome sequencing (WES), bulk and single-cell transcriptomics, spatial transcriptomics, and high-resolution spatial proteomics to resolve the spatial organization of immune evasion in EBV-positive DLBCL (**Figure 1**). Our data indicate that *PD-L1* amplification permits immune infiltration while enforcing a multilayered suppression program that combines metabolic constraints with cancer-associated fibroblast–mediated barriers, thereby limiting direct cytotoxic T-cell–tumour interactions.

**Figure 1.**
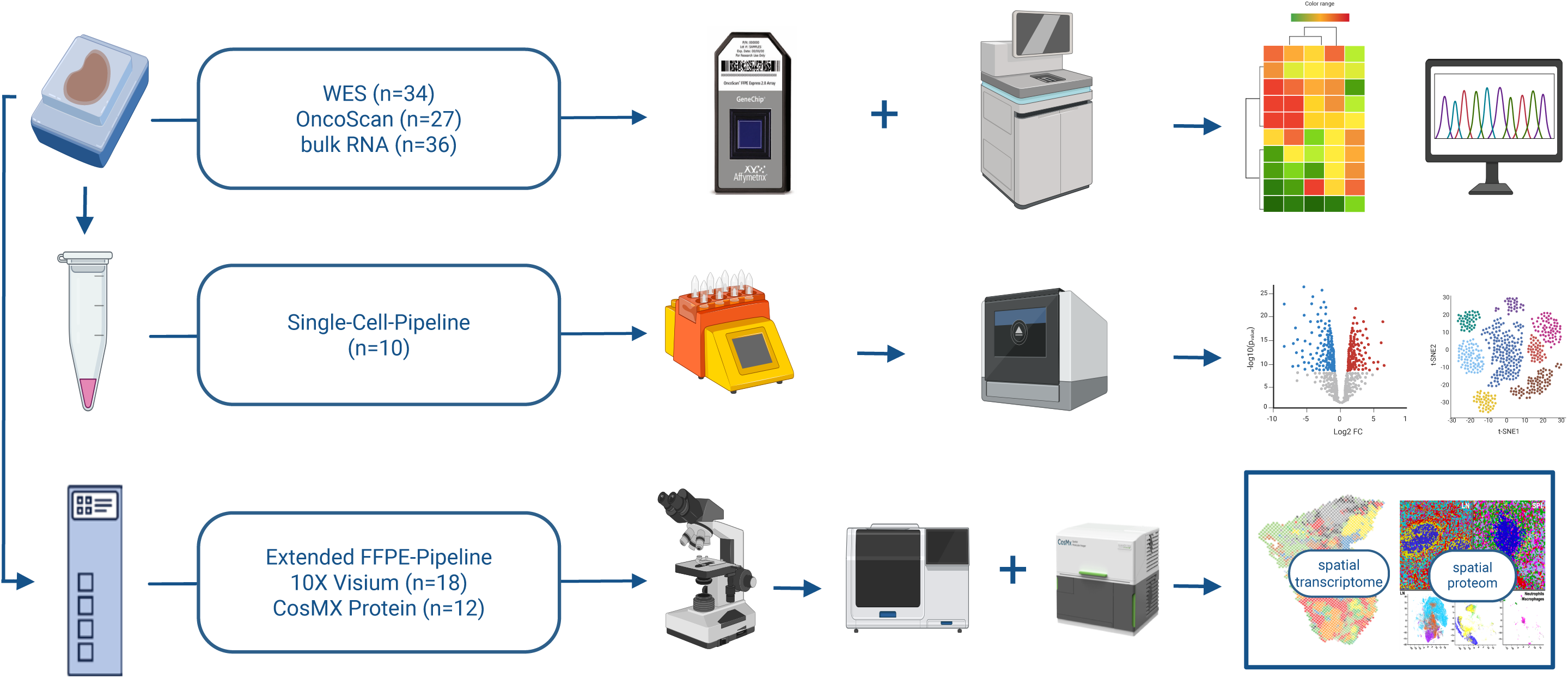
Study design and multi-omics profiling workflow. **(a)** Schematic overview of the multi-platform profiling strategy applied to EBV-positive DLBCL tumour specimens. Top arm: genomic and transcriptomic bulk profiling, comprising whole-exome sequencing (WES; n=34), OncoScan copy number arrays (n=27), and bulk RNA sequencing (n=36), yielding copy number profiles, mutational landscapes, and transcriptome-wide gene expression data. Middle arm: FFPE-based single-cell RNA sequencing pipeline (n=10 patients), yielding cell type-resolved transcriptomic profiles and differential gene expression across PD-L1 copy number groups. Bottom arm: extended FFPE spatial profiling pipeline, comprising 10x Genomics Visium spatial transcriptomics (n=18 sections) and CosMx spatial single-cell spatial proteomics with a 64-protein panel (n=12 samples), yielding spatially resolved cell type composition, ligand-receptor interactions, and tumour microenvironment architecture. Sample numbers reflect total specimens profiled. Together, these platforms enabled integrated genomic, transcriptomic, and spatial characterization of immune evasion mechanisms stratified by *PD-L1* copy number status.

## Results

### A heterogeneous genomic landscape defines EBV-positive DLBCL

We analysed 34 EBV-positive DLBCL cases that passed genomic quality control. A total of 3,447 somatic variants were identified, dominated by missense mutations (88.0%), with fewer nonsense (5.9%), splice-site (2.5%), and indel events (3.6%), and no nonstop mutations. Tumour mutational burden was low (mean 2.66, median 2.40 mutations per megabase, s.d. ±1.44; **Supplementary Table 1**).

Recurrent mutations were dispersed without a dominant driver, with frequent alterations affecting epigenetic regulators (*TET2*, *KMT2C*) and signalling pathways (*STAT3*). Mutations in canonical DLBCL subtype-defining genes (for example *MYD88*, *CD79B*, *CARD11*) were infrequent and did not define coherent molecular clusters. Cell-of-origin classification assigned most cases to the non–germinal centre B-cell subtype, with RNA-based analysis showing enrichment of activated B-cell identity alongside a substantial unclassified fraction, consistent with transcriptional heterogeneity (**Figure 2a**). *PD-L1* copy number gains at 9p24.1 were detected in 26% of cases (**Figure 2b**).

**Figure 2.**
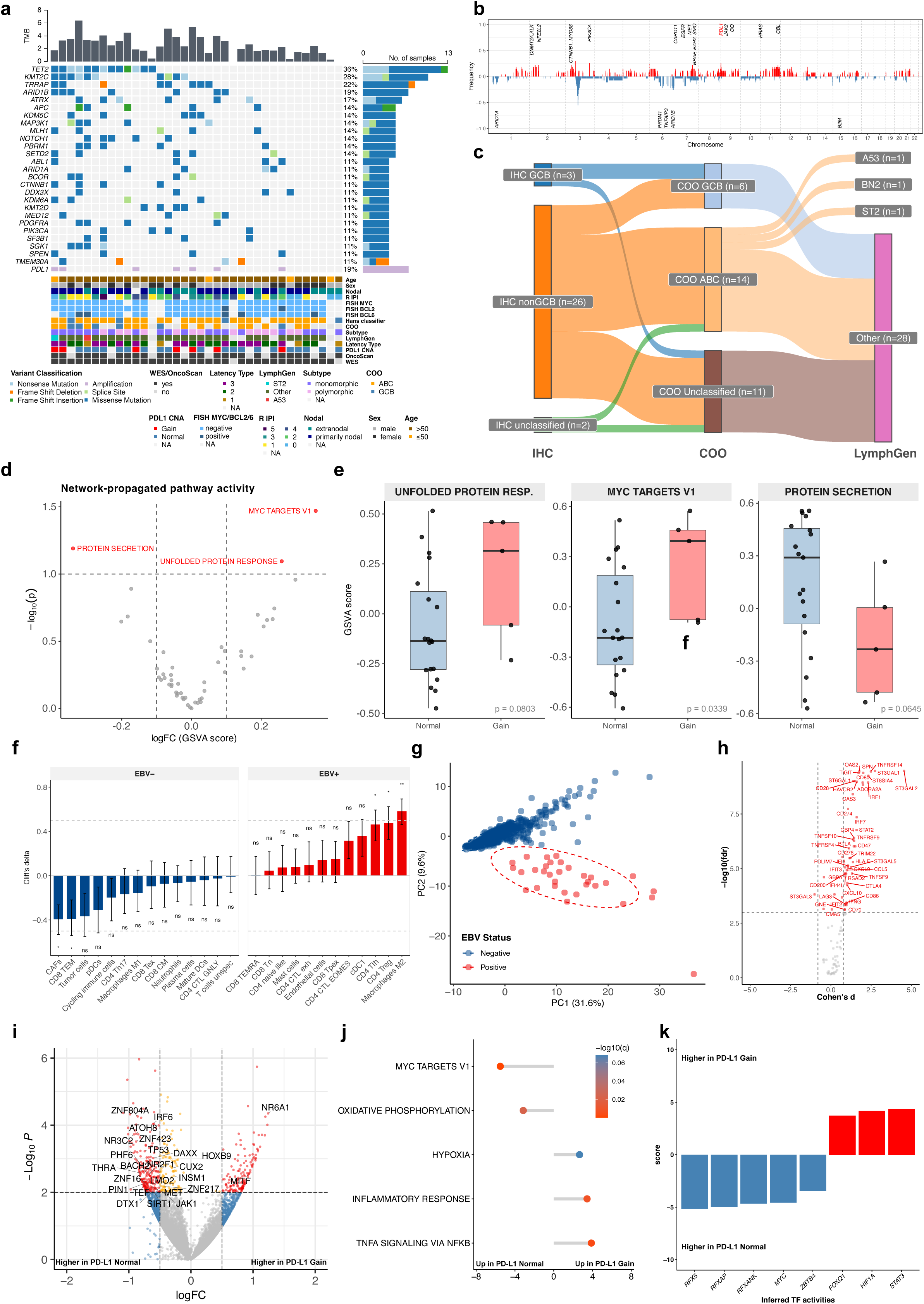
Genomic landscape and transcriptional characterization of EBV-positive DLBCL stratified by *PD-L1* copy number status. **(a)** OncoPrint showing somatic mutations and copy number alterations identified by whole-exome sequencing (WES; n=34) and OncoScan array (n=27) across the full cohort (n=36). Genes are ranked by mutation frequency. Clinical and molecular annotations are displayed below, including *PD-L1* copy number status (Gain/Normal), EBV latency type, LymphGen 2.0 classification, immunohistochemical cell-of-origin (COO) by the Hans classifier, FISH results for *MYC*, *BCL2*, and *BCL6*, revised IPI, morphology, sex, and age. **(b)** Genome-wide copy number frequency plot derived from OncoScan arrays, with amplifications (red) and deletions (blue) shown per chromosomal position. Recurrently altered loci are annotated. **(c)** Sankey plot depicting the distribution of immunohistochemical COO classification (Hans classifier; left), RNA-based COO assignment (middle), and LymphGen 2.0 subtype (right) across the cohort. **(d)** Volcano plot of network-propagated pathway activity scores comparing *PD-L1* Gain versus Normal tumours. Somatic mutations were mapped onto a protein–protein interaction network (STRING, confidence ≥ 700) and diffusion scores were computed using heat diffusion (ber_p), weighted by CADD score. Pathway activity was quantified by GSVA and differential analysis was performed with limma. Nominally significant pathways (p < 0.1) are highlighted. **(e)** Box plots of GSVA scores for the six nominally significant pathways from (d), comparing Normal and Gain tumours. P-values from limma linear models are indicated. (**f)** Cliff’s delta effect sizes for TME cell type abundance differences between EBV-positive (n=32) and EBV-negative tumours (GOYA cohort, n=553), estimated by DWLS deconvolution of bulk RNA-seq. To account for the group size imbalance, significance was assessed by Wilcoxon test over 100 subsampling iterations with balanced group sizes (n=32 per group per iteration); median p-values were FDR-corrected across cell types. Effect sizes are shown as median Cliff’s delta across iterations with 95% confidence intervals. Asterisks denote *p<0.05, **p<0.01, ***p<0.001. **(g)** Principal component analysis of bulk RNA-seq expression profiles across a curated panel of 105 immune checkpoint, sialyltransferase, and interferon-response candidate genes, coloured by EBV status. **(h)** Volcano plot showing differential expression of the 105 candidate genes between EBV-positive and EBV-negative tumours (Cohen’s d effect size versus −log₁₀ FDR). Genes significantly enriched in EBV-positive tumours are highlighted in red. **(i)** Volcano plot of bulk RNA-seq differential expression (DESeq2) comparing *PD-L1* Gain (n=6) versus Normal (n=18) EBV-positive tumours. Selected genes (CollecTRI transcription factors) with nominally significant differential expression are annotated. **(j)** Gene set enrichment analysis (GSEA; MSigDB Hallmark collection) of bulk RNA-seq data comparing *PD-L1* Gain versus Normal tumours. Normalized enrichment scores are shown as a lollipop plot, with colour indicating −log₁₀(q-value). **(k)** Transcription factor activity scores inferred by decoupleR using the DoRothEA regulon, comparing *PD-L1* Gain versus Normal EBV-positive tumours. Transcription factors are ranked by activity score; those with higher activity in Gain or Normal tumours are highlighted.

LymphGen 2.0 classification identified only 3 of 34 tumours within established subtypes, with the majority remaining unassigned, indicating limited concordance with canonical DLBCL genetic categories (**Figure 2c**).

Collectively, these data define EBV-positive DLBCL as a genetically heterogeneous entity with low mutational burden and minimal alignment with established molecular subtypes.

Exploratory subgroup analyses suggested associations between genetic alterations and phenotypic features, including enrichment of mutations in *FOXQ1, HIF1A*, and *STAT3* in *PD-L1* gain cases, *RFX5, RFXAP, RFXANK*, and *MYC* in PD-L1 wildtype cases and of *KMT2C* mutations in monomorphic tumours, as well as subtype-specific distributions across EBV latency programs. These findings were limited by sample size. To capture indirect functional effects of somatic mutations, we applied network propagation across the STRING protein–protein interaction network (v12, confidence ≥ 700), using rank-normalised CADD scores as seed weights and a regularised Laplacian kernel with permutation-based ber_p normalisation to account for topology confounding. Pathway activity was summarised using GSVA against MSigDB Hallmark gene sets and compared between *PD-L1* gain (n = 6) and copy-number normal tumours (n = 19) using limma. Nominally significant pathways (p < 0.1) identified relative enrichment of MYC Targets V1 and Unfolded Protein Response in PD-L1-gain tumours, whereas Protein Secretion was preferentially enriched in copy-number normal tumours (**Figure 2d,e**).

### EBV positivity shapes the tumour microenvironment and immune checkpoint landscape

Comparative transcriptomic analyses against EBV-negative DLBCL (n = 532; GOYA cohort ^46,47^) revealed enrichment of immunoregulatory and stromal populations, including T helper cells, regulatory T cells, and M2-polarized macrophages, in EBV-positive tumours (**Figure 2f**). Consistently, EBV-positive cases occupied an immune-enriched transcriptional space characterized by activation of interferon signalling and immune checkpoint pathways. Targeted analysis of a manually curated list of immune regulatory genes (**Supplementary Table 2**) confirmed upregulation of interferon-associated genes (*OAS2*, *IRF1*, *STAT2*) and inhibitory receptors (*TIGIT*, *PD-L1*, *LAG3*, *CTLA4, CD47, and STXGALY*), indicating concurrent immune activation and checkpoint engagement (**Figure 2g, h**).

Within the EBV-positive cohort, PD-L1 genomic gains were associated with distinct transcriptional programs. Bulk RNA-seq differential expression analysis supported widespread transcriptional differences between *PD-L1* gain and copy-number normal tumours, highlighting distinct regulatory states associated with 9p24.1 copy-number status (**Figure 2i and Supplementary Table 3**). Gene set enrichment analysis revealed distinguished inflammatory and stress-response programmes in *PD-L1* gain tumours, including TNFα signalling via NFκB, Inflammatory Response, and Hypoxia, from proliferative and metabolic programmes in copy-number normal tumours, including MYC Targets V1 and Oxidative Phosphorylation, consistent with distinct functional states associated with 9p24.1 copy-number status (**Figure 2j**). Additionally, transcription factor activities showed marked differences in relation to 9p24.1 copy-number status (q < 0.1; **Figure 2k**).

Transcriptional comparison of monomorphic (n = 19) and polymorphic (n = 13) tumours revealed subtype-specific programs, with polymorphic cases enriched for interferon-driven immune activation and STAT/IRF activity. Monomorphic tumours were dominated by MYC/E2F-associated proliferation, indicating functional heterogeneity between immune-engaged and proliferation-driven states (**Supplementary Figure S1** and **Supplementary Table 4**).

EBV latency programs further stratified transcriptional states. Latency type III tumours were enriched for interferon and TNFα/NFκB signalling alongside stress-associated pathways, whereas latency types I/II exhibited proliferation-associated programs and MYC/E2F activity, defining a continuum from proliferation-dominated to immune-activated states. Analyses involving latency type I remain exploratory due to limited sample size (**Supplementary Figure S2** and **Supplementary Table 5**).

Notably, these transcriptional differences were not accompanied by major shifts in global cellular composition, as inferred cell type abundances remained largely comparable across *PD-L1* status, histological subtypes, and latency groups, with only modest increases in immunoregulatory populations.

Together, these data define EBV-positive DLBCL as an immune-infiltrated but functionally constrained tumour ecosystem, characterized by activation of interferon-driven and checkpoint-mediated programs within a suppressive microenvironment.

### EBV latency patterns and morphological subtypes define distinct cellular and transcriptional states at single-cell resolution

Single-cell RNA sequencing was performed on 10 EBV-positive DLBCL cases, including three tumours with *PD-L1* genomic gain and seven *PD-L1* wild-type cases. A total of 112,834 high-quality nuclei were analysed enabling high-resolution dissection of tumour and microenvironmental cell states (**Figure 3a, b, Supplementary Figure S4**). Consistent with bulk deconvolution analyses, single-cell profiling confirmed that *PD-L1*-gain and *PD-L1*-wild-type tumours did not differ markedly in their overall cellular composition. Variation between individual patients was greater than *PD-L1*-status associated differences. (**Figure 3c, d**).

**Figure 3.**
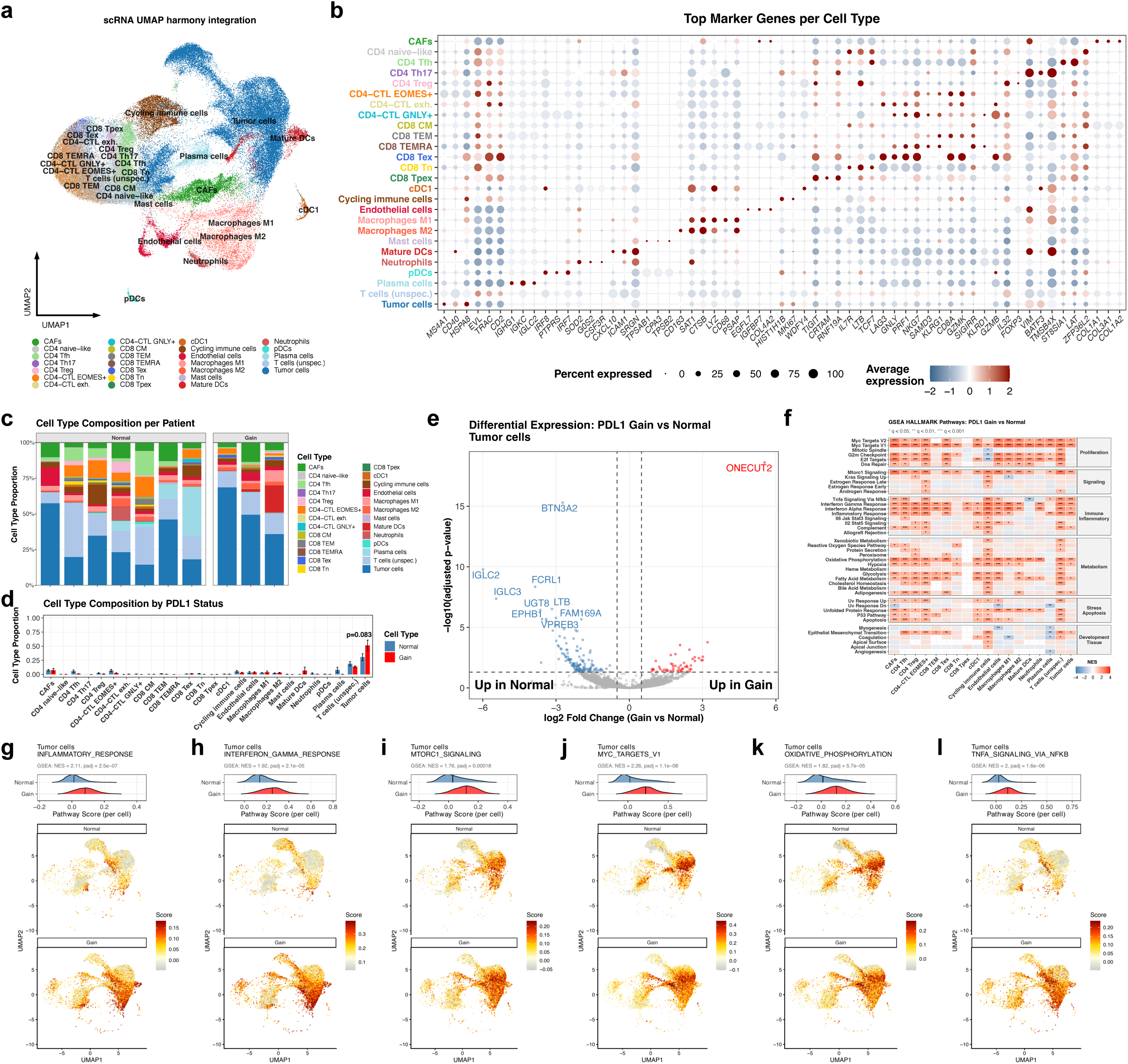
Single-cell transcriptomic landscape of EBV-positive DLBCL and *PD-L1*-associated transcriptional reprogramming. **(a)** UMAP of single-cell RNA-seq data from FFPE-derived EBV-positive DLBCL tumours (n=10 patients), integrated across patients using Harmony. Each point represents a single cell, coloured by annotated cell type. **(b)** Dot plot showing scaled average expression (colour) and detection rate (dot size) of top 5 marker genes per cell type. **(c)** Cell type composition per patient shown as stacked bar plots, ordered by *PD-L1* copy number status. **(d)** Aggregated cell type proportions stratified by *PD-L1* status (Gain/Normal), shown as grouped bar plots. P-value from permutation-based compositional testing is indicated (Brunner-Munzel test, p=0.083). **(e)** Volcano plot of pseudobulk differential gene expression (DESeq2) in tumour cells comparing *PD-L1* Gain (n=3) versus Normal (n=7) patients. Raw counts were aggregated per patient (minimum 10 cells required), genes with ≥10 counts in ≥30% of samples were retained, and log-fold change shrinkage was applied. Top differentially expressed genes are labelled. **(f)** Heatmap of gene set enrichment analysis (GSEA; MSigDB Hallmark collection) across cell types comparing *PD-L1* Gain versus Normal. Pseudobulk expression profiles per cell type were used as input; genes were ranked by signed −log₁₀(p-value) from DESeq2. Colour indicates normalized enrichment score (NES); only gene sets reaching q<0.05 in at least one cell type are shown. Pathways are grouped by biological category. Asterisks indicate *q<0.05, **q<0.01, ***q<0.001. **(g–l)** Single-cell module scores for six significantly enriched Hallmark gene sets in tumour cells: **(g)** Inflammatory Response, **(h)** Interferon Gamma Response, **(i)** mTORC1 Signalling, **(j)** MYC Targets V1, **(k)** Oxidative Phosphorylation, and **(l)** TNFα Signalling via NF-κB. Module scores were computed per cell using Seurat’s AddModuleScore with detected Hallmark gene set members as input. For each pathway, ridge plots (top) show score distributions stratified by *PD-L1* status with GSEA NES and adjusted p-value from pseudobulk analysis; UMAP projections (bottom) display per-cell scores in *PD-L1* Gain and Normal cells separately.

To resolve tumour-intrinsic transcriptional programs, malignant B cells were aggregated into pseudo-bulk profiles and stratified by EBV latency state. Differential expression analysis across latency types I–III revealed distinct transcriptional programs, with latency type III tumours exhibiting upregulation of immune-associated and interferon-responsive genes (for example, *IFIT1*, *IFI44L*, *CXCR5*), whereas latency type I/II tumours showed relative enrichment of genes linked to epithelial differentiation, inflammasome signalling, and stress-associated programs (for example, *NLRP4*, *TP63*, *MUC16*) (**Supplementary Figure S3a-c**). Pathway-level analysis further demonstrated that latency type III tumours were enriched for immune and inflammatory signalling, including interferon responses, TNFα/NFkB activation, complement, and IL6–JAK–STAT signalling, whereas latency type I/II tumours were characterized by increased activity of proliferation- and metabolism-associated pathways, including MYC targets, E2F signalling, G2M checkpoint, and oxidative phosphorylation (OXPHOS) (**Supplementary Figure S3d-t**). These findings establish latency-dependent tumour cell states spanning an immune-activated to proliferation-dominated continuum. A similar dichotomy was observed when stratifying tumours by histomorphological subtype. Pseudobulk analysis of malignant cells revealed that polymorphic EBV-positive DLBCL displayed enrichment of immune-associated transcriptional programs, whereas monomorphic tumours were dominated by proliferation and cell cycle–related pathways, including MYC and E2F target gene signatures (**Supplementary Figure S5**). Collectively, these data demonstrate that while global cellular composition remains relatively stable, EBV latency programs and morphological subtypes define distinct tumour-intrinsic transcriptional states, providing a cellular framework for the immune-engaged yet functionally constrained phenotype observed in EBV-positive DLBCL.

### *PD-L1* amplification reshapes intratumoral transcriptional programs and T-cell states toward exhaustion and restricted effector differentiation

To define the functional consequences of *PD-L1* genomic amplification at single-cell resolution, we performed pseudobulk differential expression analysis of malignant B cells and cell type–resolved pathway enrichment across *PD-L1*-gain and wild-type tumours. In malignant cells, *PD-L1* amplification was associated with distinct transcriptional reprogramming, including downregulation of B-cell lineage and antigen presentation programs (for example, *IGLC2*, *IGLC3*, *FCRL1*) (**Figure 3e**). At the pathway level, *PD-L1*-gain tumours exhibited coordinated enrichment of inflammatory and stress-response programs, including interferon gamma response, TNFα/NFκB signalling, inflammatory response, and hypoxia, alongside metabolic pathways such as OXPHOS and mTORC1 signalling (**Figure 3f-l**). These findings indicate that *PD-L1* amplification is associated with a tumour-intrinsic transcriptional state characterized by simultaneous immune activation and stress adaptation. Cell type–resolved gene set enrichment analysis indicated that these transcriptional changes extended across the tumour microenvironment, particularly within T-cell populations. CD4 Treg cells from *PD-L1*-gain tumours were enriched for MYC targets, inflammatory response, mTORC1 signalling, oxidative phosphorylation, angiogenesis, and fatty acid metabolism, consistent with a metabolically activated but functionally constrained state. CD8 TEM cells similarly showed enrichment of MYC targets and fatty acid metabolism, indicating broader metabolic remodelling of the T-cell compartment in *PD-L1*-gain tumours (**Supplementary Figure S6**). These data suggest a coordinated, multicellular transcriptional response associated with *PD-L1* amplification rather than a tumour cell–restricted effect. Spatial gradient analysis further demonstrated that these pathway alterations were spatially organized along the tumour–immune interface. In *PD-L1*-gain tumours, CD8 T cells exhibited steeper boundary-associated gradients for proliferative, metabolic and stress-response programmes, whereas Tregs showed attenuated or inverse trajectories across the same pathways, indicating compartment-specific spatial rewiring of T-cell functional states (**Supplementary Figure S7**).

Despite this broadly immune-activated transcriptional landscape, trajectory inference of CD8 T cells revealed a striking functional shift toward terminal exhaustion. Using CellRank2-based fate mapping along the CD8 Tn → Tpex → Tex axis, *PD-L1*-gain tumours exhibited significantly increased probability of terminal exhaustion compared to wild-type tumours (median 0.91 vs 0.74, Cliff’s δ = 1, Brunner–Munzel p < 0.05), accompanied by a marked reduction of the progenitor exhausted (Tpex) compartment (Cliff’s δ = 0.89, p = 0.0015) (**Supplementary Figure S8**). Consistently, UMAP projection of exhaustion trajectories demonstrated a near-uniform shift toward high exhaustion probability across all CD8 T-cell states in *PD-L1* amplified tumours, whereas wild-type tumours retained a heterogeneous distribution including a distinct low-exhaustion naïve-like population. These findings indicate that *PD-L1* amplification may accelerate differentiation along the exhaustion trajectory while depleting the self-renewing Tpex pool required for sustained anti-tumour immunity. Collectively, these results demonstrate that *PD-L1* amplification induces a paradoxical state characterized by broad immune and inflammatory activation across tumour and microenvironmental compartments, yet functionally constrains anti-tumour immunity by enforcing terminal T-cell exhaustion and limiting effector diversification. Across genomic, bulk, and single-cell analyses, *PD-L1* amplification emerged as the key determinant of tumour–immune organization, motivating a focused interrogation of its mechanistic role in shaping the EBV-positive DLBCL microenvironment.

### *PD-L1* amplification uncouples immune infiltration from tumour engagement through spatial exclusion

Spatial transcriptomic profiling of EBV-positive DLBCL samples, after quality control filtering (≥500 UMIs and ≥100 genes per spot), yielded 18 samples for analysis (**Figure 4a, b**). Cell-type composition was inferred using spatial DWLS deconvolution based on a scRNA-seq-derived signature matrix (412 genes; condition number = 68.34). Across 27 cell types, Brunner–Munzel testing with Cliff’s delta effect sizes identified selective compositional differences between *PD-L1*-gain and wild-type tumours. *PD-L1*-gain tumours were enriched for M2 macrophages (Cliff’s δ = 0.63, p = 0.020; mean 2.8% vs 9.3%) and endothelial cells (Cliff’s δ = 0.69, p = 0.020; mean 3.0% vs 5.1%), while CD8 central memory T cells were reduced (Cliff’s δ = −0.42, p = 0.134; mean 1.3% vs 0.3%) (**Figure 4c**). Spatial organization differed despite comparable overall immune abundance. In *PD-L1* wild-type tumours, cycling immune cells localized to the tumour boundary, whereas *PD-L1*-gain tumours showed depletion of these cells in tumour-adjacent regions (Δ proximity −33%), with immune cells forming peripheral aggregates. Spatial proximity analysis revealed increased immune–immune co-localization in *PD-L1*-gain tumours, including CD8 Tn–M1 macrophages (+20%), neutrophil–M1 macrophages (+20%), and CD4 Th17 interactions across multiple compartments (+20%). Suppressive associations were also increased, including M2 macrophages with neutrophils (+20%) and plasma cells (+20%), as well as CAF–M2 proximity (+20%) (**Figure 4d**). In contrast, *PD-L1* wild-type tumours showed increased tumour–immune proximity (cycling immune cells at tumour boundary: +33% relative to gain) and more distributed stromal–immune integration. Differential ligand–receptor analysis identified distinct communication architectures between groups. *PD-L1*-gain tumours were characterized by enriched inflammatory and angiogenic signalling (for example, VEGFA→NRP2, FN1→C5AR1, CCL8→ACKR1/CCR5, CD28↔CD86), whereas *PD-L1* wild-type tumours showed predominant regulatory and stromal interactions involving CAFs, CD4 T cell subsets, and endothelial cells (including TGFB1-mediated signalling and COL1A1/COL1A2→CD93 matrix interactions) (**Figure 4e, f**). Consistently, LIANA analysis revealed a more concentrated communication network in *PD-L1* gain tumours, with the highest number of significant ligand–receptor pairs observed for M1→cycling immune cells (n = 12), plasma→tumour cells (n = 10), and tumour→CD8 Tn interactions (n = 9; p < 0.01, Cliff’s δ > 0.474), whereas wild-type tumours displayed fewer but more broadly distributed interactions across CD4 T cell subsets and CAF-associated networks (**Figure 4g–k**). Together, these data indicate that *PD-L1* amplification is associated with modest compositional shifts but pronounced spatial reorganization, characterized by reduced tumour–immune contact despite preserved infiltration, consistent with spatially constrained immune engagement.

**Figure 4.**
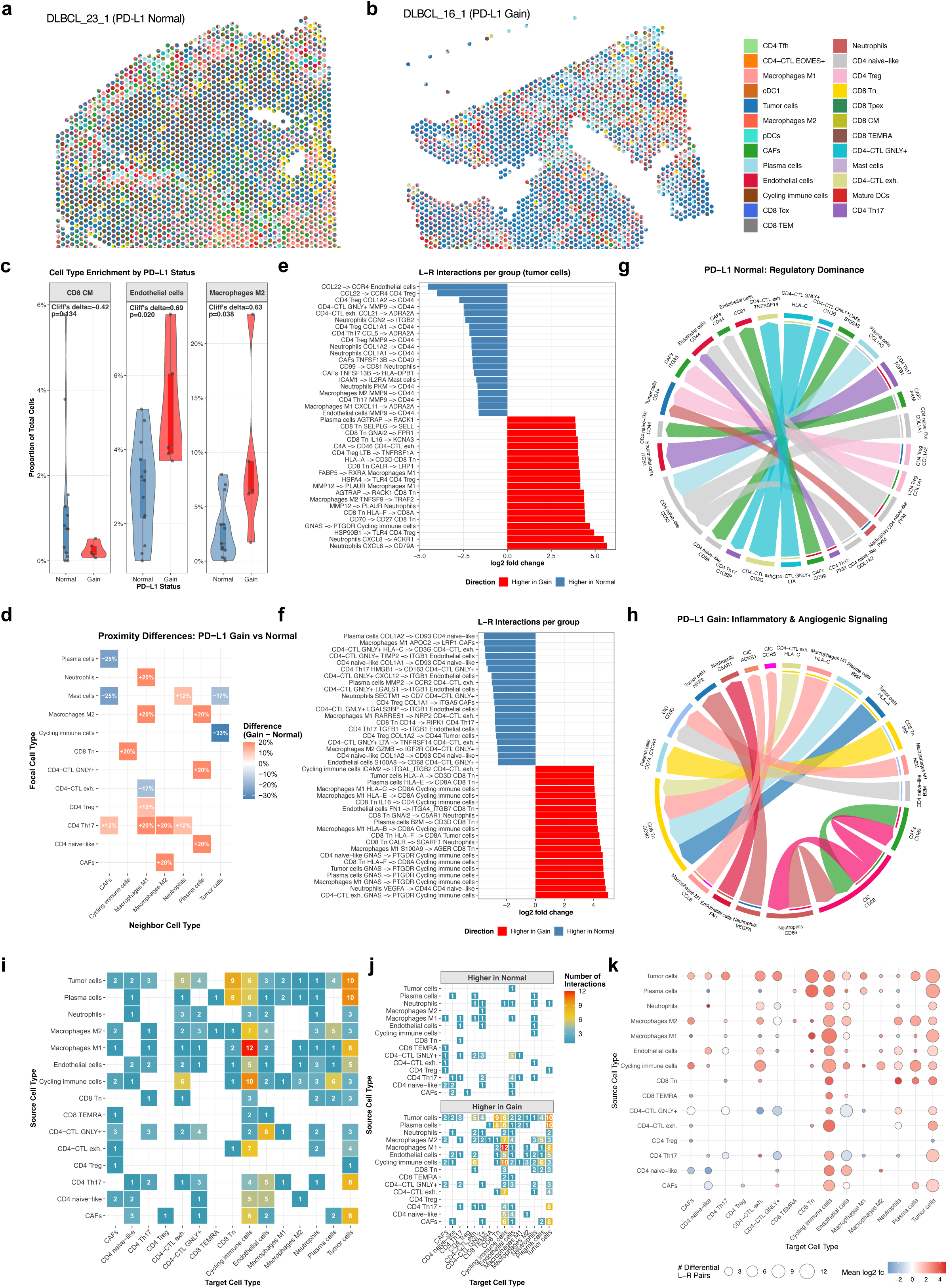
Spatial cell type composition, neighbourhood architecture, and ligand-receptor communication in EBV-positive DLBCL stratified by *PD-L1* copy number status. **(a–b)** Spatial maps of DWLS-deconvolved cell type composition for representative Visium sections from a *PD-L1* Normal tumour (DLBCL_23_1; a) and a *PD-L1* Gain tumour (DLBCL_16_1; b). Each spot is coloured by its dominant estimated cell type. **(c)** Violin plots showing the proportion of CD8 CM, Endothelial cells, and Macrophages M2 across Visium spots, comparing *PD-L1* Normal and Gain samples. Cell type proportions were estimated by DWLS deconvolution and tested using the permutation-based Brunner-Munzel test (10,000 permutations); Cliff’s delta and p-values are indicated. **(d)** Tile plot of spatial proximity differences between cell type pairs, showing the difference in the proportion of samples exhibiting significant co-occurrence (enriched proximity) between *PD-L1* Gain and Normal tumours. Proximity was assessed by k-nearest neighbour analysis (k=15) with permutation-based enrichment testing per sample; values represent Gain minus Normal proportions. **(e)** Waterfall plot of the top 20 differential ligand-receptor (L-R) interactions involving tumour cells (as source or target), comparing *PD-L1* Gain versus Normal. L-R pairs were identified by LIANA; interactions with p<0.05 and |Cliff’s delta|>0.474 are shown, ranked by log2 fold change. **(f)** Waterfall plot of the top 20 differential L-R interactions across all cell type pairs (p<0.01, |Cliff’s delta|>0.474), ranked by log2 fold change. **(g–h)** Chord diagrams summarizing the directional L-R interaction networks for the representative *PD-L1* Normal sample (g; “Regulatory Dominance”) and *PD-L1* Gain sample (h; “Inflammatory & Angiogenic Signalling”), illustrating the qualitative differences in intercellular communication architecture. **(i)** Tile heatmap showing the number of significant differential L-R pairs (p<0.01, |Cliff’s delta|>0.474) per source-target cell type combination, aggregated across the cohort. **(j)** Split tile heatmap as in (i), separately displaying interactions enriched in *PD-L1* Normal (top) and *PD-L1* Gain (bottom). **(k)** Bubble plot summarizing differential cell-cell communication by cell type pair; bubble size indicates the number of significant differential L-R pairs (p<0.01) and colour indicates the mean log2 fold change (red = higher in Gain, blue = higher in Normal).

### CAF-associated boundary enrichment coincides with reduced T cell–tumour proximity in *PD-L1*-gain tumours

To define the spatial basis of reduced tumour–immune engagement in *PD-L1* gain tumours, we quantified CAF and Treg localization relative to tumour boundaries and integrated spatial proximity with T-cell functional states.

At mesoscopic scale (100 µm), CAF boundary enrichment was increased in *PD-L1* gain tumours (Wilcoxon p = 0.030, Cliff’s δ = 0.69), with mean enrichment scores of 1.05 (+5%) versus 0.89 (−11%) in normal tumours (1.18-fold increase). This trend persisted at larger scales (p = 0.17, δ = 0.45) but was not detected at microscale resolution (p = 0.32, δ = 0.32), indicating spatially coherent CAF accumulation at the tumour interface (**Figure 5a**).

**Figure 5.**
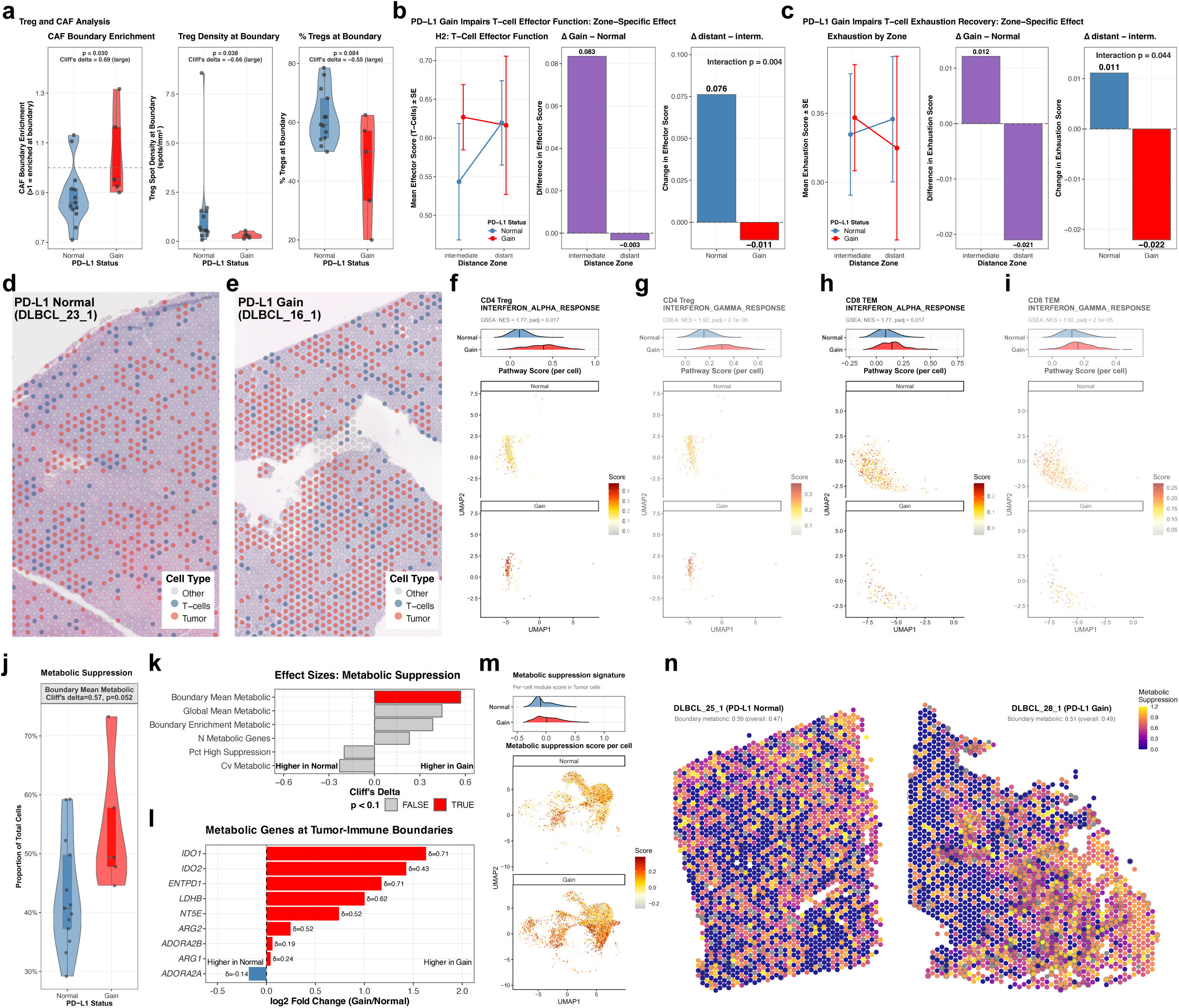
Spatial immune suppression mechanisms in EBV-positive DLBCL: CAF barriers, T-cell functional impairment, and metabolic reprogramming. **(a)** Violin plots comparing CAF boundary enrichment (ratio of CAF signal at tumour boundary vs. global; >1 indicates boundary enrichment), Treg spot density at the tumour boundary (spots/mm²), and percentage of total Tregs located at the boundary between *PD-L1* Normal and Gain Visium samples. Statistical testing was performed using the permutation-based Brunner-Munzel test; Cliff’s delta and p-values are indicated. **(b)** Zone-resolved T-cell effector function across spatial distance zones in *PD-L1* Normal and Gain tumours. Left: mean effector score (*GZMB*, *PRF1*, *IFNG*, *TNF*) ± SE per distance zone (intermediate: 100–200 µm; distant: >200 µm from tumour), estimated from spot-level linear mixed-effects models (lmer; effector_score ∼ *PD-L1* status × zone + (1|patient)), restricted to T-cell-containing spots. Centre: difference in effector score (Gain − Normal) per zone. Right: change in effector score from intermediate to distant zone per group; interaction p=0.004. **(c)** Zone-resolved T-cell exhaustion across spatial distance zones. Left: mean exhaustion score ± SE (*PDCD1*, *HAVCR2*, *LAG3*, *TIGIT*, *CTLA4*, *TOX*, *SIGLEC7*, *SIGLEC9*) per zone, estimated from spot-level generalized linear mixed-effects models (glmer; binomial family; is_exhausted ∼ *PD-L1* status × zone + (1|patient)). Centre: difference in exhaustion score (Gain − Normal) per zone. Right: change from intermediate to distant zone per group; interaction p=0.044. **(d–e)** Spatial maps of representative Visium sections from a *PD-L1* Normal tumour (DLBCL_23_1; d) and a *PD-L1* Gain tumour (DLBCL_16_1; e), with spots coloured by broad cellular category (Tumour, T-cells, Other). **(f–i)** Single-cell module scores and UMAP projections for Interferon Alpha Response (f, h) and Interferon Gamma Response (g, i) in CD4 Treg (f, g) and CD8 TEM (h, i) cells from the scRNA-seq cohort, comparing *PD-L1* Gain and Normal tumours. Ridge plots show per-cell score distributions with pseudobulk GSEA NES and adjusted p-values indicated; UMAPs display per-cell scores stratified by *PD-L1* status. **(j)** Violin plot comparing boundary mean metabolic suppression scores between *PD-L1* Normal and Gain Visium samples (Cliff’s δ=0.57, p=0.052; Brunner-Munzel test). **(k)** Cliff’s delta effect sizes for six metabolic suppression metrics derived from Visium spatial data, comparing *PD-L1* Gain versus Normal; metrics with p<0.1 (Brunner-Munzel permutation test) are highlighted. **(l)** Log2 fold change (Gain/Normal) of key metabolic suppression genes at scRNA-defined tumour-immune boundaries, with Cliff’s delta per gene indicated. Genes include adenosine pathway members (*ADORA2A*, *ADORA2B*, *ENTPD1*, *NT5E*), arginine metabolism (*ARG1*, *ARG2*), tryptophan catabolism (*IDO1*, *IDO2*), and lactate metabolism (*LDHB*). **(m)** Per-cell metabolic suppression module scores in tumour cells from the scRNA-seq cohort, shown as ridge plots and UMAP projections stratified by *PD-L1* status. **(n)** Spatial maps of metabolic suppression scores across Visium spots for representative *PD-L1* Normal (DLBCL_25_1) and *PD-L1* Gain (DLBCL_28_1) sections; boundary and global mean metabolic scores are annotated.

In contrast, boundary-associated Tregs were reduced in *PD-L1*-gain tumours. Treg density decreased from 1.40 to 0.30 spots/mm² (−78%; Wilcoxon p = 0.038, δ = −0.66), and the proportion of boundary-localized Tregs was lower (61.8% vs 44.6%; −28%; p = 0.084, δ = −0.55) (Fig. 5B, C). Global Treg–effector proximity and ratios were unchanged (all p > 0.2), indicating a boundary-restricted redistribution rather than global depletion.

Consistently, tumour–immune proximity was reduced in *PD-L1*-gain tumours, with depletion of cycling immune cells at the tumour interface (Δ −33%), despite comparable overall immune abundance. Immune cells instead showed increased immune–immune co-localization (CD8 Tn–M1 macrophages +20%, neutrophil–M1 macrophages +20%, CD4 Th17 interactions +20%) and increased CAF–M2 proximity (+20%), supporting coordinated stromal–myeloid organization. In contrast, *PD-L1* wild-type tumours showed increased tumour–immune co-localization, including enrichment of cycling immune cells at the boundary (+33% relative to gain) and higher boundary-associated Treg localization (61.8%).

To assess functional consequences, mixed-effects models were used to quantify distance-dependent changes in T-cell states across immediate, intermediate, and distant zones. In *PD-L1* wild-type tumours, T-cell effector function increased with distance from the tumour boundary (0.55 to 0.62; Δ = +0.076), whereas *PD-L1*-gain tumours lacked this recovery (0.63 to 0.62; Δ = −0.011; interaction p = 0.004, β = −0.095; main effect of distance p = 1.36×10⁻⁸). Conversely, exhaustion scores showed modest reduction with distance in wild-type tumours (0.35 to 0.34; Δ = +0.011), but increased in *PD-L1*-gain tumours (0.35 to 0.32; Δ = −0.022; interaction p = 0.044, β = −0.416; main effect p = 4.64×10⁻⁶), indicating persistent or worsening exhaustion away from the boundary (**Figure 5b, c**).

Spatial transcriptomics of representative *PD-L1*-normal and *PD-L1*-gain DLBCL samples revealed broadly comparable spatial distributions of tumour, T-cell and non-malignant compartments, indicating that *PD-L1* copy-number status is not associated with overt differences in tissue-level cellular organization (**Figure 5 d, e**). In addition, single-cell transcriptomic profiling demonstrated a consistent enrichment of interferon-responsive programs in *PD-L1*-gain tumours, with significantly increased IFN-α and IFN-γ response signatures in both CD4 Treg and CD8 TEM populations relative to *PD-L1*-normal cases (GSEA NES 1.77–1.92, adjusted P = 0.017–2.1×10⁻⁵; **Figure 5 f–i**). UMAP-based module score mapping further showed that this effect was broadly distributed across the respective T-cell compartments rather than restricted to discrete cellular subclusters, supporting a global enhancement of interferon signalling in the *PD-L1*-gain tumour microenvironment.

### Metabolic constraints reinforce immune suppression at the tumour–immune interface

To assess whether spatial immune exclusion in *PD-L1* gain tumours is accompanied by metabolic constraints, we quantified metabolic suppression signatures across tumour boundaries. *PD-L1* gain tumours exhibited increased metabolic suppression at the tumour–immune interface (mean 54.6% vs 43.1% in wild-type; Cliff’s δ = 0.57, p = 0.052), corresponding to a 1.27-fold increase specifically at boundary regions (n = 5 gain, n = 13 normal) (**Figure 5j**). Median values were similarly elevated (49.4% vs 40.7%). Global metabolic suppression showed a weaker trend (52.8% vs 45.4%; δ = 0.45, p = 0.12), whereas boundary enrichment ratios indicated preferential localization of metabolic suppression to tumour interfaces in *PD-L1*-gain tumours (boundary:global ratio 1.03 vs 0.98; δ = 0.38, p = 0.21). Spatial mapping confirmed this pattern, with *PD-L1* gain tumours displaying boundary-concentrated metabolic suppression, in contrast to diffuse distributions in wild-type tumours (representative boundary:global ratios 1.17 vs 0.74). Effect size analysis further indicated the strongest separation for boundary-specific metabolic suppression compared to global or variance-based metrics (**Figure 5 k-n**). These data indicate that metabolic suppression is spatially structured and amplified at tumour boundaries in PD-L1-gain tumours, accompanied by increased oxidative phosphorylation and amino acid catabolism, suggesting enhanced metabolic competition and nutrient deprivation at the tumour interface. Together with the observed 1.18-fold CAF boundary enrichment (p = 0.030), reduced tumour–immune contact (−23–33%), and loss of T-cell functional recovery (interaction p = 0.004), this supports a model in which a metabolically hostile boundary niche constrains T-cell fitness and reinforces spatially restricted immune–tumour engagement despite preserved overall infiltration.

### Single-cell spatial proteomics reveals contact blockade and myeloid reprogramming in *PD-L1*-gain tumours

To validate spatial organization at single-cell resolution, we performed CosMx spatial proteomics on 11 tumours (n = 8 normal, n = 3 *PD-L1*-gain; 64-plex ImmunoOncologyprotein panel), applying stringent QC and Harmony integration (**Figure 6a-c**). *PD-L1*-gain tumours showed increased T-cell proximity to tumour cells (0–50 μm: 85% vs 69%; Cliff’s δ = 0.524) and more frequent tumour nearest neighbours (34% vs 16%; ∼2-fold increase) (**Figure 6d**), yet direct CD8 T cell–tumour contact (<5 μm) was nearly absent (0.0% vs 0.6%; p = 0.020, δ = 0.667). This was accompanied by increased CAF barrier function, with a 2.6-fold higher probability of encountering CAFs in the 10–50 μm approach zone (17.5% vs 6.8%) and 2-fold enrichment of CAFs in T-cell neighbourhoods (11.3% vs 5.6%), consistent with physical intercalation between T cells and tumour cells. Mesoscopic CAF boundary enrichment was observed in both groups (ratio ∼1.0; p = 1.0), indicating that microscale positioning, rather than overall abundance, mediates functional exclusion (**Figure 6e-k**). Myeloid composition differed at single-cell resolution, with reduced total macrophage abundance in *PD-L1*-gain tumours (0.202 vs 0.080; ∼60% reduction, p = 0.36), relative enrichment of M1-like macrophages (0.072 vs 0.028; ∼2.6-fold increase), and marked depletion of M2-like macrophages (0.008 vs 0.174; ∼95% reduction, p = 0.18). Spatially, *PD-L1* wild-type tumours exhibited boundary-associated M2 enrichment (0–10 μm: 81.7%), whereas *PD-L1*-gain tumours showed uniformly reduced M2 proportions across all distances (0–10 μm: 58.4%; 10–20 μm: 56.6%; 20–50 μm: 55.0%; 50–100 μm: 50.3%; >100 μm: 50.8%) and loss of polarization gradients (M1:M2 ∼1:4 vs ∼1:1). Macrophage polarization was spatially organized in normal tumours but disrupted in *PD-L1*-gain tumours, with persistent M1 skewing at tumour boundaries and absence of coordinated distal M2 enrichment. These data suggest spatial compartmentalization of myeloid populations, with transcriptional profiling capturing distal stromal regions enriched in M2 macrophages, whereas single-cell proteomics highlights tumour-proximal depletion and architectural remodelling (**Figure 6l**). Interpretation is limited by panel composition (absence of TCF7, PRF1, IFNG, TNF, CD25), but core exhaustion and lineage markers enable robust assessment of spatial organization. Collectively, these findings confirm a multi-scale immune evasion mechanism in *PD-L1*-gain tumours, characterized by preserved infiltration and tumour proximity but CAF-mediated blockade of direct T cell–tumour contact, accompanied by disrupted myeloid organization and loss of spatial polarization.

**Figure 6.**
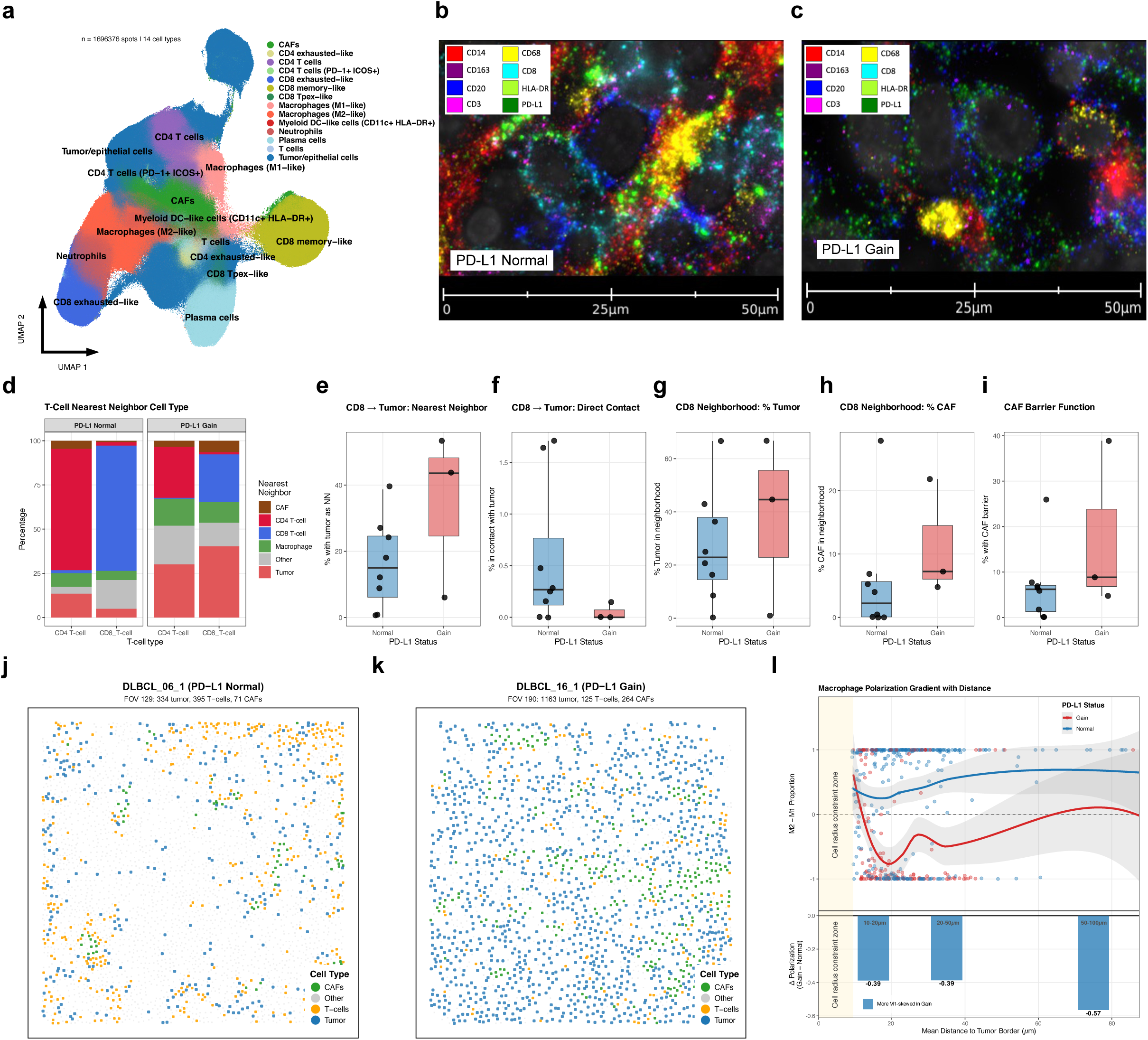
Spatial single-cell spatial proteomics by CosMx reveals microscale CAF-mediated T-cell exclusion and macrophage polarization gradients in *PD-L1* Gain tumours. **(a)** UMAP of 1,696,376 single cells profiled by CosMx spatial single-cell spatial proteomics (64-protein panel) across n=10 EBV-positive DLBCL samples (Normal n=7, Gain n=3), annotated into 14 cell types. **(b,c)** Multiplex protein fluorescence images of representative CosMx FOVs from a *PD-L1* Normal (b) and *PD-L1* Gain (c) tumour, showing co-staining for CD14, CD163, CD8, CD20, CD3, HLA-DR, and PD-L1. Scale bar = 50 µm. **(g)** Stacked bar plots showing the distribution of nearest-neighbour cell types for CD4 and CD8 T-cells separately, in *PD-L1* Normal and Gain tumours. Proportions are computed at patient level by averaging across FOVs per sample. **(d–i)** Patient-level summary metrics for CD8 T-cell spatial relationships comparing *PD-L1* Normal and Gain tumours, computed by aggregating FOV-level measurements per sample: percentage of CD8 T-cells with a tumour cell as their nearest neighbour (e), percentage of CD8 T-cells in direct contact with tumour cells (f), percentage of tumour cells in the CD8 T-cell neighbourhood (g), percentage of CAF cells in the CD8 T-cell neighbourhood (h), and percentage of CD8 T-cells with a CAF as the nearest neighbour between T-cell and tumour (CAF barrier function; j). All comparisons tested using the Brunner-Munzel test with Cliff’s delta as effect size measure. (l**B** Macrophage polarization gradient as a function of distance from the tumour border. (Top) M2−M1 proportion (proportion of M2-like minus proportion of M1-like macrophages among all macrophages per FOV) plotted against mean distance to the tumour border (µm). Each point represents one field of view (FOV; restricted to FOVs with ≥5% tumour content and ≥5 macrophages); lines show LOESS-smoothed trends with 95% confidence intervals, stratified by *PD-L1* copy-number status (Gain, red; Normal, blue). The shaded yellow zone indicates the cell radius constraint zone, where centroid-to-centroid distances below ∼9.5 µm reflect the physical exclusion imposed by non-overlapping cell segmentation masks rather than true spatial separation. Distances were calculated within individual FOVs as the Euclidean distance from each macrophage centroid to its nearest tumour cell centroid using physical coordinates (µm; pixel-to-µm conversion: 0.18 µm/pixel). (Bottom) Δ Polarization (Gain − Normal) per distance bin, quantifying the between-group difference in mean M2−M1 proportion at each spatial zone. Values were computed from cell-level distance assignments aggregated to FOV then sample level prior to group averaging, avoiding mean-of-means distortion introduced by FOV-level mean distance binning. Negative values (blue) indicate greater M1-skewing in *PD-L1* Gain; positive values (red) indicate greater M2-skewing. **(j,k)** Representative CosMx field-of-view (FOV) spatial maps from a *PD-L1* Normal tumour (DLBCL_06_1, FOV 129; i) and a *PD-L1* Gain tumour (DLBCL_16_1, FOV 190; k), with cells coloured by broad category (Tumour, T-cells, CAFs, Other). Cell counts per category are indicated in the panel titles.

**Figure 7.**
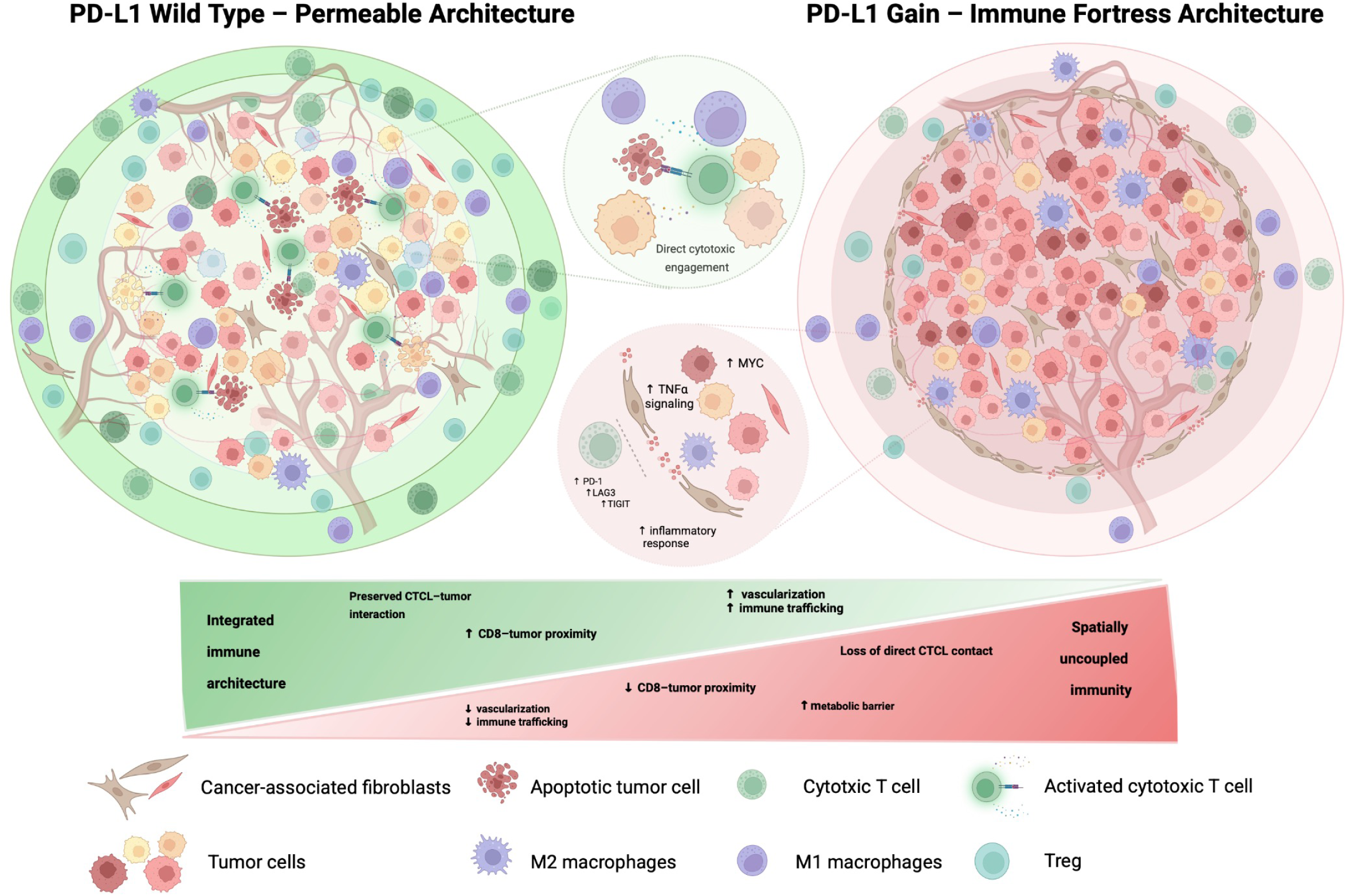
Integrative model of *PD-L1* copy number-dependent immune architecture in EBV-positive DLBCL. Schematic comparison of the tumour microenvironment organization in *PD-L1* Normal (left, “Permeable Architecture”) and *PD-L1* Gain (right, “Immune Fortress Architecture”) tumours. In *PD-L1* Normal tumours, cytotoxic T-cells access the tumour core and engage in direct cytotoxic contact with tumour cells, supported by preserved vascularization and immune trafficking, and an M1-skewed macrophage polarization. In *PD-L1* Gain tumours, a spatially organized suppressive barrier emerges: cancer-associated fibroblasts (CAFs) form a stromal boundary that physically excludes cytotoxic T-cells from direct tumour contact, while regulatory T-cells are depleted at the interface. Tumour-intrinsic transcriptional reprogramming is characterized by upregulation of MYC targets, TNFα signalling, and inflammatory response pathways, accompanied by co-expression of multiple immune checkpoints (PD-1, LAG3, TIGIT) on exhausted T-cells. Reduced vascularization and impaired immune trafficking further limit effector T-cell infiltration, and a metabolic suppression barrier restricts T-cell function in the tumour-proximal zone. Together, these mechanisms converge on a state of spatially uncoupled immunity, in which T-cells are present in the periphery but functionally and physically barred from tumour engagement. The gradient bar (bottom) summarizes the transition from integrated immune architecture to spatially uncoupled immunity across the *PD-L1* copy number spectrum. Cell type legend is shown below.

## Discussion

In this study, we demonstrate that EBV-positive DLBCL is characterized by a primed immunosuppressive environment not through immune exclusion but through a spatially organized decoupling of immune infiltration from immune engagement, in which T cells remain abundant and proximal to tumour cells yet fail to establish productive cytotoxic interactions. This state is enforced by a multilayered program integrating PD-L1–driven signalling, fibroblast-mediated physical barriers, metabolic suppression at tumour interfaces, and terminal T-cell exhaustion, collectively defining EBV-positive DLBCL as an immune-infiltrated but spatially disengaged tumour ecosystem.

Dysfunctional T cells have been reported to accumulate in close proximity to EBV-positive malignant B cells across multiple contexts, suggesting that spatially proximal but ineffective immune responses may be a recurring feature of EBV-driven lymphomagenesis. EBV-positive DLBCL has thus far been primarily characterized through small cohorts. Compared to EBV-negative disease, these characterizations were primarily conducted without considering the heterogeneity within the EBV-positive group itself ^44,45,48,49^. Our integrative analysis extends these observations by resolving the spatial organization and functional consequences of tumour–immune interactions across genomic, transcriptional, and single-cell spatial scales.

EBV-driven oncogenic programs establish a baseline immune-evasive state by promoting proliferation and impairing immune recognition through EBNA2-mediated regulatory circuits, including suppression of ICOSL via miR-24 and upregulation of PD-L1 through miR-34a downregulation, thereby reducing tumour immunogenicity while maintaining pro-proliferative c-MYC activity ^50–52^. Our data suggest that these virus-driven mechanisms create a permissive immunosuppressive environment upon which additional genetic events act.

In this context, *PD-L1* amplification emerges as a cooperative reinforcement of an already active immune evasion axis. EBV-driven signalling establishes baseline *PD-L1* expression through pathways such as NFκB and JAK–STAT, while secondary genomic amplification likely acts as a state transition event, raising this signal beyond a functional threshold required for stable, spatially organized immune suppression, as observed in other tumour entities including gastric adenocarcinoma ^35,53^.

Beyond its canonical role as an immune checkpoint ligand, PD-L1 may thus function as a higher-order regulator of tumour–immune ecosystem organization, coordinating transcriptional programs, intercellular communication, and immune cell states across multiple compartments^54,55^.

Consistent with this model, *PD-L1* amplification functionally uncouples immune infiltration from immune engagement, with T cells remaining spatially proximal to tumour cells yet failing to establish effective cytotoxic interactions. This phenotype parallels tumour contexts in which immune cells are present but functionally restrained within the microenvironment ^56,57^. This suggests that immune evasion is governed not only by immune cell abundance or checkpoint expression, but by the spatial organization of tumour–immune interactions that determines whether immune recognition can be translated into effective anti-tumour activity. Other DLBCL subtypes, particularly primary CNS and testicular lymphomas, also frequently harbour *PD-L1* amplification, suggesting that immune evasion in these entities arises through active immunoediting rather than mere immune privilege. Consistent with this, recent analyses have demonstrated that these lymphomas are characterized by substantial immune cell infiltration organized in complex, spatially heterogeneous niches composed of diverse cellular mixtures, paralleling our observations of immune-rich yet functionally constrained tumour ecosystems^44,58–60^.

EBV-positive DLBCL is therefore not adequately captured by the classical dichotomy of immune-desert and immune-excluded tumours, but instead represents an immune-infiltrated yet functionally disengaged state defined by spatially constrained immune activity. This extends current frameworks of tumour immune phenotypes toward a model in which immune presence and immune function are uncoupled at the level of tissue architecture ^61^.

Mechanistically, our data support a multilayered model of immune evasion integrating CAF-mediated physical barriers, loss of direct tumour–T cell contact, boundary-associated metabolic suppression, and enforced terminal T-cell exhaustion. This convergence of structural, metabolic, and functional constraints is consistent with observations in other tumour entities, such as pancreatic ductal adenocarcinoma and triple-negative breast cancer, where stromal exclusion, metabolic reprogramming, and progressive T-cell dysfunction act in concert to limit effective anti-tumour immunity ^56,62,63^. The apparent discrepancy between spatial transcriptomics and single-cell spatial proteomics is not discordant but orthogonal, reflecting differences in spatial resolution, with transcriptomic approaches capturing broader stromal compartments and imaging resolving tumour-proximal niches. Integrated analysis supports a model of spatial compartmentalization in which immunosuppressive programs are regionally distributed and differentially manifested across tumour ecosystems, consistent with recent multi-modal studies in other solid tumours demonstrating hierarchical organization of fibroblast and myeloid niches ^64,65^. The myeloid compartment does not conform to a simple polarization shift but instead reflects a disruption of spatial organization, with depletion of M2-like macrophages in tumour-proximal niches despite their enrichment at the tissue level. This spatial compartmentalization indicates that immune suppression is structurally orchestrated through coordinated stromal–myeloid interactions rather than driven by uniform cell-intrinsic states, consistent with observations in other tumour contexts such as pancreatic ductal adenocarcinoma and hepatocellular carcinoma, where myeloid cells occupy spatially defined niches that regulate tumour–immune interactions ^66,67^.

Consistent with prior reports, PD-1 blockade in combination with chemoimmunotherapy has demonstrated high activity in EBV-positive DLBCL. Our data further suggest that co-targeting additional immune checkpoints such as LAG3 and CTLA4, both significantly upregulated in our analyses, may enhance therapeutic efficacy by addressing complementary mechanisms of immune restraint ^68–70^.

Our results support an evolving framework for DLBCL classification in which genetic alterations, viral programs, and tumour microenvironmental architecture jointly define biologically and clinically relevant subgroups. The spatial features identified here—including fibroblast-mediated exclusion and immune cell distribution patterns—represent candidate phenotypes that, once validated in larger cohorts and across independent settings, could be inferred from routine histopathology using machine learning, enabling scalable assessment of tumour–immune architecture in clinical practice ^71,72^.

Several limitations should be acknowledged. First, cohort sizes within individual modalities, particularly single-cell and spatial analyses, remain moderate given the rarity of EBV-positive DLBCL, and larger studies will be required to resolve inter-patient heterogeneity and refine subtype-specific effects. Second, the cross-sectional design precludes inference of temporal dynamics linking EBV-driven signalling, *PD-L1* amplification, and spatial immune organization. Third, spatial deconvolution and ligand–receptor analyses rely on model-based approaches that may incompletely capture rare or transient cell states, and functional conclusions are largely based on correlative transcriptional and spatial readouts rather than direct perturbation. Finally, while our data suggest that tumour–immune architecture may be inferable from routine histopathology, this remains to be validated in independent clinical cohorts.

Together, these findings define EBV-positive DLBCL as a spatially organized immune-evasive ecosystem in which viral signalling and *PD-L1* amplification cooperatively rewire tumour–immune interactions. By linking genetic and microenvironmental architecture to immune dysfunction, our work provides a rationale for spatially informed therapeutic strategies.

## Methods

### Human research ethics approval

This study was approved by the Institutional Review Board of the University of Lübeck (18-356) using archival samples and a waiver of informed consent. Clinical baseline characteristics of study participants are provided in **Supplementary Table 6**. No compensation was provided to participants.

### Case selection and clinicopathological assessment

For this retrospective analysis, we reviewed our institutional archive for cases of histologically confirmed EBV-positive DLBCL between January 2008 and December 2021. Histopathological work-up was performed as described and diagnosis was confirmed following the 5^th^ edition WHO classification of hematopoietic and lymphoid tumours and the ICC criteria and yielded 34 primary diagnostic EBV-positive DLBCL samples from 34 patients for which sufficient formalin-fixed paraffin-embedded (FFPE) tissue samples were available for subsequent molecular analysis ^7,18,73^. No samples of relapsed and/or refractory disease were included.

### Delineation of the molecular composition of the study cohort FISH for MYC, BCL2, and BCL6

Chromosomal breakpoints were analysed by means of FISH using dual colour break apart probes for 8q24 (*MYC*), 18q21 (*BCL2*) and 3q27 (*BCL6*) (Abott Vysis Des Plaines, IL, USA) according to the manufacturer’s instructions and as described ^74^.

### Extraction of nucleic acids, whole exome and RNA sequencing and Detection of Somatic Copy Number Alterations

Molecular characteristics were assessed as recently described ^73,75^. Genomic tumour DNA as well as tumour RNA were each extracted from three FFPE tissue sections of 5µm thickness using Maxwell® RSC DNA FFPE kit and Maxwell® RSC RNA FFPE kit (both Promega).

Whole-exome sequencing (WES) was performed employing a hybrid capture approach with the Agilent SureSelect XT Human All Exon V6 library preparation kit (Agilent Technologies) followed by massively parallel sequencing of enriched exonic sequences on a NovaSeq platform (Illumina).

The raw sequencing data in FASTQ format was processed using the NFORE workflow (NEXTFLOW v24.04.2) SAREK (v3.4.2) for variant calling against the GRCh38 reference genome ^76,77^. Initially, reads were trimmed for adapter sequences and quality using FASTP (v0.23.4), followed by alignment with BWA MEM2 (v2.2.1) ^78,79^. Subsequent steps included mate-pair information correction, removal of PCR duplicates, and base quality recalibration utilizing PICARD TOOLS, GATK (v4.5.0.0), and dbSNP v146 ^80,81^. Finally, variant calling was executed in tumour-only mode using MUTECT2 with GNOMAD (r2.1.1) as the reference for known germline variants ^82,83^. Quality control metrics in accordance with the recommendation by Dreval *et al.* are summarized in **Supplementary Table 7** ^84^. A brief graphical representation of the bioinformatics workflow for WES data processing is provided in **Supplementary Figure 9**.

The transcriptional profile of EBV-positive DLBCL was investigated by RNA sequencing (RNA-seq). Ribo-Zero Magnetic Kit (Human/Mouse/Rat) (Illumina) was used for rRNA depletion and library preparation was performed, employing NEBNext® UltraT Directional RNA Library Prep Kit (New England BioLabs).

Somatic copy number alterations were detected using OncoScan CNV assays (ThermoFisher). Raw data (*CEL* files) were processed using the EACON package (v0.4.0) with ASCAT^85,86^ as segmentation algorithm. *L2R* files in CBS format were used as input for GISTIC (v2.0.23)^87^ to identify regions that are significantly amplified or deleted across all samples (confidence level 0.90, focal length cutoff 0.50, q-value threshold 0.1). *PD-L1* copy number status at 9p24.1 was derived from OncoScan ASCAT segmentation data. Samples were classified as *PD-L1* gain if the ASCAT-estimated log₂ ratio at the *CD274* locus exceeded 0, corresponding to any copy number increase above diploid; samples with log₂ ratio ≤ 0 were classified as copy-number normal. A threshold of log₂ ratio > 0 was applied rather than the conventional > 0.2 cutoff to account for the modest signal ratios (range 0.08–0.82) expected when single-copy gains occur in FFPE tissue with residual normal cell admixture.

### Filtering of FFPE-derived artifacts, FLAGS, SNPs and low-quality variant calls

Variants were left-aligned (GATK LEFTALIGNANDTRIMVARIANTS) and only variants with a PASS filter flag were kept for variant annotation using VARIANT EFFECT PREDICTOR (VEP v111, GRCh38; adding CADD v1.6, dbNSFP v4.1a, and GNOMAD r3.0 as additional annotations) ^88–90^. Next, annotations were converted into *MAF* format using VCF2MAF (V1.6.21) (DOI:10.5281/ZENODO.593251); coverage was extracted directly from the vcf INFO field. Afterward, low-quality samples (low average median coverage) were removed as well as potential germline variants in the data using GATKs 1000g Panel of Normals. The top 20 frequently mutated genes (FLAGS)^91^ were removed from further analysis. Somatic variants with a frequency ≥ 0.1% in 1000 genomes, GNOMAD, or ExAC were removed, and the resulting somatic variants were screened for FFPE artifacts (detected using SOBDetector v1.0.4^92^ and excerno v0.1.0^93^), which were excluded as well. The remaining somatic variants were filtered in a two-step procedure. (1) High-impact variants (CADD score > 20 or likely pathogenic according to AlphaMissense) in candidate genes (see below) were filtered as such that minimum coverage of 10, minimum alternative coverage of 4, and minimum variant allele frequency of 5% were required. Candidate genes defined as follows: (a) listed as tumour suppressor or oncogene according to Vogelstein et al. ^94^, (b) genes from the Lymphgen algorithm, (c) DLBCL genes described by Chapuy *et al.*, (d) MHC genes according to Harmonizome (v3.0; StandardizedValue ≥ 1) ^29,58,95^. (2) For low-impact variants (CADD score ≤ 20) were filtered as follows: minimum coverage of 50, minimum alternative allele coverage of 5, minimum variant allele frequency of 10%, variants were required to have a COSMIC or dbSNP ID. Both variant lists (high and low-impact) were merged for downstream analysis.

### Somatic mutation network diffusion and pathway enrichment

To capture indirect functional consequences of somatic mutations beyond directly mutated genes, network propagation was performed across the STRING protein–protein interaction network (v12.0, combined confidence score ≥ 700). Highly connected hub proteins (degree ≥ 900, including ubiquitin) were removed prior to analysis. Somatic variants were pre-filtered to retain high-confidence calls (CADD PHRED ≥ 20, tumour allele frequency > 5%). For each sample, mutated genes were assigned seed weights equal to the maximum rank-normalised CADD score across all variants affecting that gene, prioritising the most damaging variant per locus. Scores were propagated using a pre-computed regularised Laplacian kernel and normalised with the permutation-based ber_p statistic (1,000 permutations; diffuStats v1.32.0) to account for network topology confounding. Sample-level diffusion score matrices were summarised into pathway activity estimates using GSVA (v2.6.0; kcdf = “Gaussian”, minSize = 4, maxSize = 500) against MSigDB Hallmark gene sets. Pathway activity was compared between *PD-L1*-gain (n = 6) and copy-number normal (n = 19) tumours using limma (v3.68.1) with robust empirical Bayes moderation (eBayes, robust = TRUE); pathways with nominal p < 0.1 are reported as hypothesis-generating given the limited sample size (**Supplementary Table 8**).

### Transcriptome Data Quantification and Analysis

RNA-seq data (fastq files) was processed using the NF-CORE/RNASEQ pipeline (v3.13.2). FASTP trimmed reads were pseudo-aligned to GRCh38 using SALMON (v1.10.1) ^96^. Fusions (BCL2/6) were detected using RNAFUSION (v3.0.1; NFCORE) against GRCh38 as reference genome; FASTP was used to remove low-quality bases from the sequencing data ^97,98^.

Differential gene expression analysis was performed using DESEQ2 (v1.52.0)^99^.To reduce variance from *log_2_* fold-changes, a heavy-tailed Cauchy prior distribution for effect sizes was used (shrinkage of *log_2_* fold-changes) ^100^. Genes with fewer than 10 counts summed across all samples were excluded prior to testing. Gene set enrichment analysis against MSigDB HALLMARK gene sets (MSIGDF R package v2026.1) on DESEQ2 estimated shrinked *log_2_* fold-changes was performed using GAGE (v2.62.0); results with a corrected q-value below 0.1 (Benjamini-Hochberg corrected) were considered significant.

To assess transcriptional differences relative to EBV-negative DLBCL, EBV-positive cases were compared against the GOYA cohort (ENA accession number PRJNA518119; n = 532 EBV-negative cases)^46^. For targeted expression comparisons, a curated panel of 114 immune regulatory genes was analysed, comprising T-cell immune checkpoints, myeloid immune checkpoints, sialic acid pathway components and sialyltransferases, EBV immune response genes, antigen-presenting cell co-stimulatory molecules, and T-cell receptor signalling mediators, as well as *SPN* were included based on prior reports of differential expression in EBV-positive lymphomas. TPM values were used for all candidate gene comparisons. To account for the substantial size imbalance, group-level comparisons were performed using 100-iteration Wilcoxon rank-sum subsampling, randomly drawing n = 32 EBV-negative samples per iteration; results were summarised as median effect size (Cliff’s δ) and empirical p-values across iterations.

Transcription factor activity was inferred using decoupleR (v2.17.0) with the univariate linear model method (run_ulm) and the CollecTRI regulon as the prior network. Per-sample activity scores were estimated from variance-stabilising transformation (VST)-normalised DESeq2 counts. Contrast-level TF activity was additionally estimated by applying run_ulm to DESeq2 t-statistics, with Benjamini–Hochberg correction applied to the resulting p-values.

Bulk tumour immune composition was estimated by DWLS (Dampened Weighted Least Squares; v0.1.0)^101^ deconvolution for the EBV-positive versus EBV-negative comparison. A cell-type signature matrix was derived from the scRNA-seq reference dataset using Seurat FindAllMarkers (v5.5.0; Wilcoxon rank-sum test, one-vs-all; Benjamini–Hochberg-adjusted p < 0.05, average log₂ fold-change > 0), retaining the top 20 markers per cell type ranked by average log₂ fold-change. Average expression per cell type was computed on scaled data and used as the signature matrix. DWLS was applied per sample after trimming the signature and bulk matrices to their common gene set. Cell-type proportion estimates were compared between EBV-positive and EBV-negative groups using 100-iteration subsampling Wilcoxon rank-sum tests with Cliff’s δ as effect size, with Benjamini–Hochberg correction applied to median p-values across iterations.

### Molecular Subtyping

Molecular subtype classification was performed using LymphGen 2.0. Somatic variants were converted to MAF format and lifted over from GRCh38 to hg19 coordinates using liftOver prior to input generation. BCL2 and BCL6 translocation status was determined as described above (fusionreport composite score > 0.1). Input files were prepared using the LGenIC utility with full copy number data (CGH.class = 0), and classification was performed using the LymphGen for R implementation (Predict9), testing all six subtypes (BN2, EZB, MCD, N1, ST2, A53). Samples with no subtype confidence exceeding 0.5 were classified as Other.

Classification into ABC/GCB subgroups (cell of origin) was performed using expression profiles of previously described candidate genes for GCB (*ITPKB*, *MME*, *BCL6*, *MYBL1*, *DENND3*, *NEK6*, *LMO2*, *LRMP*, *SERPINA9*) and ABC (*SH3BP5*, *IRF4*, *PIM1*, *ENTPD1*, *BLNK*, *CCND2*, *ETV6*, *FUT8*, *BMF*, *IL16*, *PTPN1*) ^102^. Briefly, expression profiles (counts) were quantile normalized, *log_2_* transformed, and *z*-normalized across genes. For each subtype, a score was computed for each sample *i,* and gene *j* follows

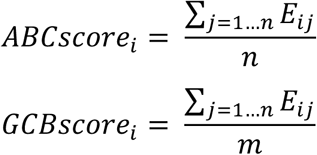

*E_ij_* was defined as the expression of gene *j* in sample *i*, *n* was defined as the number of ABC genes with expression > 0, and *m* was defined as the number of GCB genes with expression > 0. The subtype score was defined as

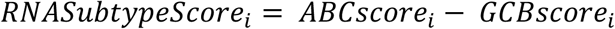

A sample with an *RNASubtypeScore* above 0.25 was considered as ABC-subtype, below -0.25 as GCB-subtype, respectively.

### Chromium Single Cell Gene Expression Flex

Cells and Nuclei were isolated from two 50-µm FFPE scrolls following the demonstrated protocol for cell isolation from FFPE tissue sections for Chromium Fixed RNA Profiling (CG000632). Library preparation and sequencing were performed according to the Chromium Fixed RNA Profiling for Multiplexed Samples User Guide (CG000527, revision D).

### Single-cell RNA sequencing preprocessing, integration, and cell type annotation

#### Data processing and alignment

Raw FASTQ files were demultiplexed and aligned to the human reference genome using Cell Ranger version 7.2.0 (10x Genomics). Samples were aligned to the GRCh38-2024 reference genome (refdata-gex-GRCh38-2024) using probe set *Chromium_Human_Transcriptome_Probe_Set_v1.0_GRCh38-2020-A*. Libraries were processed using a dedicated configuration file specifying the corresponding sample barcodes. Cell Ranger output per-sample count matrices were used for further analyses.

Subsequent analyses were performed using R and the Seurat package (version 5.0.3). Count matrices for each sample were loaded using the *Read10X()* function. Empty droplets were identified and removed using *DropletUtils* (v1.22.0). Retained barcodes were used to construct Seurat objects, requiring a minimum of 3 cells per gene. Doublet detection was performed using s*cDblFinder* (version 1.16.0). Cells classified as doublets were removed, retaining only singlets for downstream analysis. Quality control filtering was applied per sample based on sample-specific thresholds for mitochondrial percentage, number of detected genes (*nFeature_RNA*), and total UMI count (*nCount_RNA*), as summarised in **Supplementary Table 9** (**Supplementary Figure 10**) Cells exceeding the maximum mitochondrial percentage or falling outside the defined count and feature ranges were removed, where a total of 112,834 cells remained.Seurat objects from all 10 samples were merged into a single object and processed as follows: data were log-normalised (scale factor 10,000) and the top 3,000 highly variable features were identified using the variance-stabilising transformation (VST) method, scaled and principal component analysis (PCA) was performed on the top 30 principal component. Integration across samples was performed using Harmony, implemented via the *IntegrateLayers* function in Seurat (*HarmonyIntegration* method), correcting for sample-of-origin as the integration variable. Clustering was performed on the Harmony-corrected embedding using *FindNeighbors()* (30 dimensions) and *FindClusters* with the with default parameters and resolution of 0.1. Uniform manifold approximation projection (UMAP) embeddings were computed on 30 dimensions of the Harmony reduction for visualisation. After integration, layers were rejoined using *JoinLayers*.

Major cell populations were manually annotated using canonical marker genes curated from the literature. Annotated lineages included T cells, B/tumour cells, macrophages, neutrophils, plasma cells, cancer-associated fibroblasts, endothelial cells, dendritic cells, and mast cells. To further characterize macrophage heterogeneity, cells annotated as macrophages were extracted and classified into polarization states using literature-derived markers associated with M1-like and M2-like phenotypes Specifically, CD68 and CD163 were used as representative markers of M1-like and M2-like macrophages, respectively, based on a previously published EBV-positive DLBCL phenotyping strategy^45^. Module scores for each marker set were calculated using Seurat’s *AddModuleScore* function. An M2 proportion score was subsequently computed as the ratio of the M2 module score to the sum of the M1 and M2 module scores. Macrophages were classified as M1 when the M1 score exceeded the M2 score, and as M2 when the M2 score exceeded the M1 score. Cells exhibiting high expression of both signatures (M1 score > 0.5 and M2 score > 0.5) were classified as M0/Hybrid, while cells not meeting these criteria were labelled as Uncertain. M0/Hybrid and Uncertain macrophages were excluded from downstream analyses (**Supplementary Figure 11**).

### Pseudobulk differential expression and gene set enrichment (scRNA-seq)

Pseudobulk count matrices were generated by summing raw counts across all cells of each cell type per patient. For *PD-L1* status comparisons, cell types were included if at least 10 cells per patient were available; patients with fewer than 10 cells for a given cell type were excluded, and cell types with fewer than five eligible patients were skipped. For latency-type pairwise comparisons, thresholds were relaxed to ≥5 cells per patient and ≥4 eligible patients total, to accommodate the smaller subgroup sizes. For morphological subtype comparisons (monomorphic vs polymorphic), the same thresholds as for the *PD-L1* comparison were applied.

Differential expression was performed using DESeq2 (v1.52.0) per cell type independently, with design formulae ∼PDL1_CNA, ∼LatencyType, or ∼Subtype as appropriate. For *PD-L1* and subtype comparisons, genes were retained if they showed ≥10 counts in at least 30% of samples and cell types with fewer than 100 genes after filtering were excluded. For latency comparisons, thresholds were relaxed to ≥5 counts in ≥25% of samples and a minimum of 50 retained genes. Log₂ fold-change shrinkage was applied using a normal prior (lfcShrink, type = “normal”). Pairwise latency-type contrasts were performed for all three combinations (type I vs II, type I vs III, type II vs III). Differentially expressed genes were defined by Benjamini–Hochberg-adjusted p < 0.05.

Gene set enrichment analysis was performed per cell type using fgsea (v1.38.0; fgseaMultilevel) against MSigDB Hallmark gene sets (msigdf v2026.1; minSize = 15, maxSize = 500). Genes were ranked by signed −log₁₀(p-value) derived from DESeq2 results (sign(log₂FC) × −log_10_(p-value)). Gene sets with Benjamini–Hochberg-adjusted p < 0.05 were considered significant. For visualisation, Hallmark gene sets were grouped into thematic categories covering all 50 pathways.

Cell type composition was compared between *PD-L1*-gain and copy-number normal tumours at the patient level using Brunner–Munzel permutation tests, requiring a minimum of four patients per comparison.

To visualise enrichment of significant pathways at single-cell resolution, per-cell module scores were computed within individual cell types using Seurat AddModuleScore for Hallmark gene sets reaching adjusted p < 0.001 in the corresponding pseudobulk enrichment analysis (restricted to gene sets with ≥5 detected genes), and displayed as density ridge plots stratified by *PD-L1* status and as UMAP overlays.

### Metabolic suppression analysis (scRNA-seq)

To assess whether metabolic immune suppression is enriched at tumour–immune boundaries in *PD-L1*-gain tumours, a boundary activation score and a metabolic suppression score were computed per cell using Seurat AddModuleScore. The boundary signature comprised *IFNG*, *IFNGR1*, *STAT1*, *IRF1*, *CXCL9*, *CXCL10*, *CD8A*, *CD8B*, *PRF1*, *GZMB*, *GZMA*, *CD274*, *PDCD1*, *HAVCR2*, and *LAG3*. The metabolic suppression signature comprised *ENTPD1*, *NT5E*, *ADORA2A*, *ADORA2B*, *LDHA*, *LDHB*, *IDO1*, *IDO2*, *ARG1*, and *ARG2*. Boundary cells were defined as those in the upper quartile (≥75th percentile) of the boundary score; core cells as those in the lower quartile (≤25th percentile). The primary analysis tested whether boundary enrichment of metabolic suppression differed between *PD-L1*-gain and copy-number normal tumours using a linear mixed-effects model (lmer: Metabolic_Score ∼ PDL1_gain × boundary + (1 | Patient); lmerTest). For conservative patient-level validation, boundary enrichment was quantified per patient as the difference in mean metabolic score between boundary and core cells and compared between groups using Brunner–Munzel permutation tests (10,000 permutations) with Cliff’s δ as effect size. Individual metabolic gene expression at boundary cells was additionally tested at the patient level using the same framework, with Benjamini–Hochberg correction applied across genes.

### CD8 T-cell exhaustion trajectory inference

To assess fate commitment along the CD8 T-cell exhaustion axis, trajectory inference was performed using CellRank2^103^. T cells and tumour cells were extracted from the integrated Seurat object and exported to AnnData format with raw counts, Harmony-integrated UMAP coordinates, and cell-level metadata. Analysis was restricted to the exhaustion axis subset (CD8 Tn, CD8 Tpex, CD8 Tex). Count data were normalised to 10,000 counts per cell, log₁p-transformed, and 3,000 highly variable genes were identified (flavor = “seurat_v3”). PCA (50 components) and a neighbour graph (30 PCs, k = 12 neighbours) were recomputed on this subset independently of the Harmony integration, to capture T-cell-intrinsic differentiation axes. A gene expression-based pseudotime was defined per cell as the score of exhaustion marker genes (*PDCD1*, *LAG3*, *HAVCR2*, *TIGIT*, *TOX*, *ENTPD1*) minus the score of naive marker genes (*TCF7*, *CCR7*, *SELL*, *LEF1*), computed using scanpy score_genes and min–max normalised to [0, 1]. A CellRank2 PseudotimeKernel was constructed from this pseudotime and a transition matrix computed toward higher pseudotime values. Terminal and intermediate macrostates were identified using the GPCCA estimator (Schur decomposition, n_components = 10, n_states = 4), with CD8 Tn excluded from terminal states. CD8 Tex was resolved into two macrostates (Tex_1 and Tex_2); their fate probabilities were summed into a single CD8 Tex fate probability per cell. Fate probabilities were aggregated to patient level as the weighted mean across CD8 T-cell subtypes, weighted by cell count per subtype. The Tpex compartment size was quantified as the Tpex / (Tpex + Tex) fraction per patient. Patient-level fate probabilities and Tpex fractions were compared between *PD-L1*-gain and copy-number normal tumours using Brunner–Munzel tests with Cliff’s δ as effect size.

### Visium Spatial Gene Expression

Following re-embedding of 5x5 mm tissue segments according to histologically confirmed regions of interest carrying characteristic features of the tumour FFPE serial sections (5 µm) were placed on 10X Genomics Visium slides and stained with H&E following the Demonstrated Protocol Visium CytAssist Spatial Gene Expression for FFPE — Deparaffinization, H&E Staining, Imaging & Decrosslinking. Samples were processed and sequenced following the Visium Spatial Gene Expression Reagent Kits User Guide.

### Visium spatial transcriptomics data processing

Raw sequencing data were processed using Space Ranger (v3.1.0; 10x Genomics) with the GRCh38-2020-A reference genome and the Visium Human Transcriptome Probe Set v1.0 (GRCh38-2020-A). The spaceranger count pipeline was run per sample in FFPE mode with image reorientation enabled (--reorient-images=true), using slide ID and capture area coordinates provided per sample. Alignment and feature–barcode matrix generation were performed on a high-performance computing cluster. BAM file generation was disabled to reduce storage requirements.

### Visium spatial transcriptomics preprocessing and quality control

Space Ranger output data were loaded per sample and subjected to ambient RNA decontamination using SpotClean (v1.14.0), with a maximum of 50 iterations, candidate radius of 20 spots, and gene cutoff of 0.01. Decontaminated count matrices were converted to Seurat objects and filtered to retain spots with ≥500 UMI counts and ≥100 detected genes; genes were retained if they showed ≥100 total UMI counts and were detected in ≥5 spots. Artefact-associated gene sets — mitochondrial, heat-shock protein, ribosomal, and dissociation-related genes, as defined by Xue et al.^104^ — were quantified per spot for quality monitoring but not used as hard filters. Sample-level quality was assessed using the ambient RNA contamination (ARC) score; samples with concurrent low feature counts (median < 400 genes/spot) and low UMI counts (median < 1,000 UMIs/spot), or with ≥2 quality flag criteria, were classified as poor quality and excluded. A total of 18 samples passed quality control and were taken forward (in total 4 samples excluded) (**Supplementary Table 10**).

### Cell type deconvolution (Visium)

Cell type composition was inferred per spot using DWLS (Dampened Weighted Least Squares) deconvolution implemented in Giotto (v4.2.2). A cell-type signature matrix was derived from the scRNA-seq reference using Giotto’s scran-based one-vs-all marker detection (findMarkers_one_vs_all), retaining the top 20 markers per cell type ranked by effect size (412 genes total; condition number = 68.34). Average expression per cell type was computed on normalised data and used as the signature matrix input for spatialDWLS. Cell-type proportion estimates were compared between *PD-L1*-gain and copy-number normal groups using Brunner–Munzel tests with Cliff’s δ as effect size, with Benjamini–Hochberg correction.

### Tumour boundary zone definition and spatial gradient analysis (Visium)

Tumour and immune spot identity was derived from DWLS deconvolution, with spots assigned to the dominant cell type by maximum estimated proportion. Distance from each non-tumour spot to the nearest tumour spot was calculated using k-nearest neighbour search (RANN, k = 1). Spots were assigned to four distance zones relative to the tumour boundary: immediate (0–50 µm), proximal (50–100 µm), intermediate (100–200 µm), and distant (>200 µm). CAF boundary enrichment was quantified at three spatial scales — micro (25th percentile of non-tumour distances), meso (50th percentile), and macro (75th percentile) — as the ratio of mean CAF score within the boundary zone to the global mean CAF score. Treg density (spots/mm²) and the proportion of boundary-localised Tregs were quantified at the meso scale. CAF and Treg boundary metrics were compared between *PD-L1*-gain and copy-number normal tumours using Wilcoxon rank-sum tests with Cliff’s δ as effect size and Benjamini–Hochberg correction. Spatial cell-type co-occurrence was assessed per sample using a k-nearest neighbour graph (k = 15; dbscan::kNN) constructed from spot coordinates, with each spot assigned its dominant cell type by maximum DWLS proportion. For each focal–neighbour cell-type pair, observed co-occurrence within neighbourhoods was compared against the expected count under random neighbour composition; an enrichment ratio (observed/expected) and a z-score ((observed − expected)/√expected) were computed, with pairs requiring an expected count ≥5. Normal-approximation p-values were FDR-corrected, and a pair was considered significantly enriched at FDR < 0.05. Self-interactions (same cell type; reflecting clustering) and cross-interactions (different cell types; reflecting proximity) were analysed separately. Between-group differences in proximity were summarised per cell-type pair as the difference in the proportion of samples exhibiting significant enriched cross-interaction between *PD-L1*-gain and copy-number normal tumours.

T-cell effector function and exhaustion were scored per spot as the mean log-normalised expression of their respective signature genes. The effector signature comprised *GZMB*, *PRF1*, *IFNG*, and *TNF*; the exhaustion signature comprised *PDCD1*, *HAVCR2*, *LAG3*, *TIGIT*, *CTLA4*, *TOX*, *SIGLEC7*, and *SIGLEC9*. Each non-tumour spot was assigned to a distance zone relative to the nearest tumour spot (immediate 0–50 µm, proximal 50–100 µm, intermediate 100–200 µm, distant >200 µm), with distances computed by nearest-neighbour search (RANN::nn2, k = 1). Mixed-effects modelling was restricted to the intermediate (100–200 µm) and distant (>200 µm) zones, as the immediate (0–50 µm) and proximal (50–100 µm) zones contained too few T-cell-containing spots for stable estimation. Distance-dependent changes in effector function were modelled using a linear mixed-effects model (lme4/lmerTest) with *PD-L1* status, zone, and their interaction as fixed effects and patient as a random intercept (effector_score ∼ *PD-L1* status × zone + (1 | sample_id)); the *PD-L1* × zone interaction was the primary test term, with p-values obtained by Satterthwaite approximation. T-cell exhaustion was modelled at the spot level as the proportion of exhausted T cells (defined as CD8 Tex and exhausted CD4 cytotoxic T cells) using a generalised linear mixed-effects model with binomial error and a logit link (glmer: is_exhausted ∼ *PD-L1* status × zone + (1 | sample_id)). Estimated marginal means and pairwise group contrasts per zone were obtained using emmeans.

Metabolic immune suppression was quantified per Visium spot using a signature comprising *ENTPD1* (CD39), *NT5E* (CD73), *ADORA2A*, *IDO1*, and *ARG1*. Genes were retained if detected in ≥2% of spots, requiring a minimum of two genes; the metabolic suppression score per spot was computed as the mean log-normalised expression across available genes. For each sample, a global mean score, a boundary mean score (spots within 150 µm of the nearest tumour spot), and a boundary enrichment ratio (boundary mean / global mean) were derived, along with the proportion of high-suppression spots (≥75th percentile) and the coefficient of variation. These six metrics were compared between *PD-L1*-gain and copy-number normal tumours using Brunner–Munzel permutation tests (10,000 permutations) with Cliff’s δ as effect size and Benjamini–Hochberg correction.

### Ligand–receptor communication analysis (Visium)

Cell–cell communication was inferred per sample using LIANA (v0.1.14) with the Consensus ligand–receptor resource. Three scoring methods were applied in parallel — NATMI, Connectome, and SingleCellSignalR — and interaction scores were aggregated using liana_aggregate to produce a consensus rank. Significant interactions were defined by aggregate rank ≤ 0.01. For between-group comparisons (*PD-L1* Gain vs Normal), aggregate ranks per ligand–receptor pair were compared using Wilcoxon rank-sum tests with Cliff’s δ as effect size, and results were summarised by sender–receiver cell type pair.

### Single Cell Spatial Proteomics

Spatial single-cell proteomic profiling of 5-µm FFPE lymphoma biopsy sections was performed using the CosMx™ Spatial Molecular Imager (SMI; NanoString/Bruker) with the 64-plex CosMx™ Human Immuno-Oncology Protein Panel (NanoString/Bruker, #121500010), following the manufacturer’s instructions.

### CosMx spatial single-cell spatial proteomics preprocessing and quality control

Raw CosMX data were exported from the NanoString AtoMx platform as per-slide expression matrices, cell metadata, FOV position files, and cell polygon files. Sample identities were assigned to individual cells based on FOV ranges using slide-specific mapping files. Non-biological markers were removed from the protein panel before analysis, comprising five channel controls (Channel-CD3, Channel-CD45, Channel-DNA, Channel-Membrane, Channel-PanCK) and two isotype controls (Ms IgG1, Rb IgG), leaving 57 biological proteins for analysis. Global spatial coordinates were calculated per cell by combining FOV-level positions with local pixel coordinates using a conversion factor of 0.18 µm/pixel.

Quality control filtering was applied using multiple criteria: cells were required to pass all built-in NanoString QC flags (area, negative probe, high expression, and low expression); cell area was restricted to 8–200 µm² to exclude debris and doublets; aspect ratio was restricted to 0.4–2.5 to remove severely distorted cells; and mean negative probe signal was required to be < 25 to minimise background (**Supplementary Table 11**). Filtered cells were assembled into Seurat objects per slide. Protein expression was normalised using centred log-ratio (CLR) transformation per cell (NormalizeData, normalization.method = “CLR”).

### Batch correction and dimensionality reduction (CosMx)

Data from both slides were merged with slide-specific cell identifiers. PCA was performed on CLR-normalised protein expression across all proteins (npcs = 30). Based on elbow plot inspection, the first 10 principal components were selected for Harmony batch correction (RunHarmony; group.by.vars = “SampleID”, dims.use = 1:10, max_iter = 20), correcting for technical slide-to-slide variation while preserving biological heterogeneity. UMAP embedding was computed on Harmony-corrected dimensions (dims = 1:10). All subsequent distance calculations were performed within individual FOVs using FOV-local coordinates to prevent biologically invalid cross-FOV distance measurements.

### Cell type annotation (CosMx)

Unsupervised clustering was performed using the Leiden algorithm (FindNeighbors on Harmony reduction, dims = 1:10; FindClusters, resolution = 0.8, algorithm = 4, seed = 123). Cluster marker proteins were identified by Wilcoxon rank-sum one-vs-all testing (FindAllMarkers; min.pct = 0.25, log₂FC threshold = 0.25). Clusters were annotated by computing a geometric overlap score between the top 10 cluster markers (avg_log₂FC > 0.35) and curated protein signatures for each expected cell type; clusters with overlap score ≥ 0.15 were assigned the best-matching label. Clusters failing initial annotation were re-clustered independently at the same resolution and re-annotated with relaxed thresholds (avg_log₂FC > 0.32, min overlap score = 0.10). Tumour cells were defined as clusters with CD19 or CD20 as differential markers; plasma cells were classified as tumour microenvironment rather than tumour. Final cell type labels reflect conservative manuscript-ready nomenclature acknowledging the limitations of protein-based annotation (e.g., “CD8 exhausted-like”, “Macrophages (M1-like)”).

### Proximity, contact, and neighbourhood analysis (CosMx)

For each cell, the nearest neighbour of any cell type was identified within the same FOV using k-nearest neighbour search (RANN::nn2, k = 2; the first neighbour being the cell itself). Five spatial metrics were computed per CD8 T cell per FOV and aggregated to patient level by averaging across FOVs: (1) the proportion of CD8 T cells whose nearest neighbour is a tumour cell; (2) the proportion of CD8 T cells in direct contact with a tumour cell (nearest-neighbour distance < 5 µm); (3) the proportion of tumour cells in the CD8 T-cell neighbourhood (within 20 µm radius); (4) the proportion of CAFs in the CD8 T-cell neighbourhood (within 20 µm radius); and (5) the proportion of CD8 T cells in the 10–50 µm approach zone from the tumour boundary whose nearest neighbour is a CAF (CAF barrier function). Neighbourhood composition was computed using a radius-based search (RANN::nn2, searchtype = “radius”, radius = 20 µm; maximum 100 neighbours per cell). FOVs with fewer than 5 cells were excluded. All five metrics were compared between *PD-L1*-gain and copy-number normal tumours at patient level using Brunner–Munzel tests with Cliff’s δ as effect size.

Macrophage polarisation gradients were computed as the M2-like minus M1-like macrophage proportion among all macrophages per FOV, plotted as a function of mean distance to the tumour boundary, restricted to FOVs with ≥5% tumour content and ≥5 macrophages. Trends were fitted using LOESS smoothing weighted by the number of macrophages per FOV, stratified by *PD-L1* copy-number status.

Macrophage compositional differences were assessed from CosMx cell-type annotations. Per FOV, the total macrophage proportion and the proportions of M1-like and M2-like macrophages were computed and aggregated to patient level by averaging across FOVs. Patient-level proportions were compared between *PD-L1*-gain and copy-number normal tumours using Wilcoxon rank-sum tests. To account for within-patient FOV correlation, FOV-level proportions were additionally modelled using linear mixed-effects models with *PD-L1* status as a fixed effect and patient as a random intercept (prop ∼ *PD-L1* status + (1 | SampleID)), fitted separately for total, M1-like, and M2-like macrophages.

### Statistical analysis

If not reported otherwise, statistical analysis was performed using R (v4.6.0), and p-values were corrected using Benjamini-Hochberg correction. The following R packages were used: TIDYVERSE (v2.0.0)^105^ for data handling and plotting; MAFTOOLS (v2.28.0)^106^ to summarize, analyse, and visualize whole exome sequencing data; visualizations were generated using ggplot2 (v4.0.3) and patchwork (v1.3.2); ENHANCEDVOLCANO (v1.30.0)^107^ was used to produce plot volcano plots. For two-group comparisons with small and unbalanced sample sizes, the Brunner–Munzel test was used as a nonparametric analogue of Welch’s t-test that does not assume equal variances or identical distribution shapes under the null and is valid without requiring equal group sizes [CITE^108^]; effect sizes are reported as Cliff’s δ throughout (using EFFSIZE package v0.8.1). Cliff’s δ was interpreted using the following thresholds: negligible < 0.147, small < 0.33, medium < 0.474, large ≥ 0.474. Where repeated measurements were nested within patients, linear mixed-effects models were fitted using lme4 (v2.0-1) and lmerTest (v3.2-1) with patient as a random intercept; p-values were obtained by Satterthwaite approximation.

### Pseudonymization

The processing of personal and baseline clinical data for this study was performed pseudonymously by using a case ID (**Supplementary Table 7**). Due to pseudonymization data backtracking specific to the individual for non-members of the study group is nearly impossible. Only the initiators of the study (NG, AK) have access to a file that is separately stored (password-protected) containing the details on pseudonymization.

## Data Availability

Raw FASTQ files have been deposited in the European Genome-phenome Archive (EGA) under accession numbers EGAD50000002793 (whole-exome sequencing), EGAD50000002792 (single-cell RNA sequencing), and EGAD50000002791 (Visium spatial transcriptomics). OncoScan array data have been deposited in the Gene Expression Omnibus (GEO) under accession number GSE335860. Processed single-cell RNA sequencing (Seurat object), Visium spatial transcriptomics (Space Ranger output and Seurat objects), and CosMx spatial proteomics data (AtoMx export and Seurat object) have been deposited at Zenodo under DOI 10.5281/zenodo.20796627.

## Ethics approval and consent to participate

This retrospective study was approved by the ethics committee of the University of Lübeck (reference-no 18-356) and conducted in accordance with the declaration of Helsinki. Patients have provided written informed consent regarding routine diagnostic and academic assessment, including genomic studies of their biopsy specimens alongside transfer of their clinical data.

## Funding

This work was supported by generous funding by the Damp-Stiftung through a project grant (NG). HB, AK, and SD acknowledge support by the BMBF project OUTLIVE-CRC (FKZ 01KD2103A).

## Notes

Conflict of interest: TR: Former employee of Johnson & Johnson (08/2022–04/2025). Since 05/2025 employed at the University Hospital Schleswig-Holstein. No further conflicts of interest

### Competing Interest Statement

TR: Former employee of Johnson & Johnson (08/2022-04/2025). Since 05/2025 employed at the University Hospital Schleswig-Holstein. No further conflicts of interest

### Author Declarations

This study was approved by the Institutional Review Board of the University of Luebeck (18-356) using archival samples and a waiver of informed consent.

